# Immune Profiling after treatment with an Anti-CD19 Chimeric Antigen Receptor T Cell Therapy in Treatment-Refractory Progressive Multiple Sclerosis

**DOI:** 10.64898/2026.09.11.26361992

**Authors:** Sasha Gupta, Madhav R. Seshadri, Ravi Dandekar, Leonie Müller-Jensen, Krista McCutcheon, Leslie A. Scarffe, ShiLu Vanasupa, Megumi Sunahara, Fumie Hayashi, Colette Caspar, Robin Lincoln, Naomi Okinishi, Nancy Thomas, Samantha Shenoy, Samantha Zylberman, Camille Fouassier, Ebtesam Hassan, Sam Klauer, Jenai Wilmoth, Joseph J. Sabatino, Aaron Bodansky, Ahmed Abdelhak, Stephen L. Hauser, Michael R. Wilson, Bruce A. C. Cree

## Abstract

Treatment targeting progressive multiple sclerosis (MS) have limited efficacy, as central nervous system (CNS) resident B cells are resistant to peripheral B cell depletion by anti-CD20 monoclonal antibodies. CD19-targeted chimeric antigen receptor (CAR)-T cells offer the promise to deplete B cells more extensively in both the peripheral and CNS compartments. Two adult participants with treatment-refractory progressive MS were treated with 0.33×10^8 autologous anti-CD19 CAR-T cells (mivocabtagene autoleucel, miv-cel) intravenously after a preconditioning regimen. Participant 1 (50’s year old female, EDSS 6) had no cytokine release syndrome (CRS) or immune effector cell-associated neurotoxicity syndrome (ICANS). Participant 2 (60’s year old female, EDSS 4.5) had grade 1 CRS and no ICANS. Expansion of CAR-T cells in the cerebrospinal fluid (CSF) at Day 14 was noted in both participants, remarkably with sustained disappearance of disease-associated oligoclonal band (OCB) or normalization of IgG index despite peripheral naive B cell reconstitution. MRI and clinical outcomes showed sustained stability at Week 48. Serologic studies showed a decline in anti-Epstein Barr Virus seropositivity. Single cell myeloid and proteomic signatures showed a decline in proinflammatory profiles.

In the 2 subjects with progressive MS reported here, low dose anti-CD19 CAR-T cell therapy (miv-cel) was safe, penetrated the CSF effectively, reduced intrathecal humoral activity, modulated peripheral immune tone, and appeared to stabilize EDSS scores. Overall, these results support the continued investigation of CAR-T cell therapy in MS.

**One Sentence Summary:** Anti-CD19 CAR-T cell was safe in two progressive MS patients with CSF penetration, OCB/IgG index resolution, and improved inflammatory background.

## INTRODUCTION

Many people living with multiple sclerosis (MS) will ultimately develop a progressive form of the disease, characterized by a steadily increasing symptom burden which may include physical, cognitive, and other deficits. Despite the U.S. Food and Drug Administration’s (FDA) approval of siponimod for active secondary progressive MS (SPMS) [1], ocrelizumab for primary progressive MS [2], and the European Medicines Agency approving tolebrutinib for SPMS [3], these therapies have modest benefits in slowing disability accumulation. There is a clear and pressing unmet need for more effective treatments for patients with progressive MS, especially those with worsening symptoms despite treatment with disease modifying therapies (treatment-refractory MS).

B cells appear to be integral to the pathogenesis of MS. B cell depletion with anti-CD20 monoclonal antibody (mAb) therapy strongly suppresses relapsing disease activity [4]. B cells are involved in cytokine secretion and antigen presentation to T cells [5] as well as being the source of antibody production, including clonally expanded immunoglobulin G (IgG) oligoclonal bands (OCBs) detectable in the cerebrospinal fluid (CSF) of about 90% of MS patients [6]. Ectopic meningeal lymphoid aggregates – a central nervous system (CNS) manifestation of tertiary lymphoid organ formation recognized to occur in other organ systems in various autoimmune diseases and malignancies – are present in MS, especially in the later stages of disease. These meningeal follicles are associated with adjacent cortical demyelination and neuronal injury and harbor B cells [7–10].

Despite effectively suppressing relapsing disease activity, anti-CD20 mAbs do not eradicate OCBs or attenuate ectopic meningeal lymphoid aggregates [11, 12]. Peripherally administered anti-CD20 monoclonal antibody therapies may not gain adequate access to the CNS to efficiently deplete resident B cells, especially in patients with “compartmentalized” inflammation with an intact blood-brain barrier. Further, currently approved anti-CD20 mAbs utilize several mechanisms of action to deplete B cells including CD20 cross-linking to induce apoptosis, complement dependent cytotoxicity (CDC), antibody dependent cellular cytotoxicity (ADCC) or phagocytosis (ADCP). While abundant in peripheral blood, complement, natural killer cells and macrophages are far less abundant in the CNS, and their scarcity is likely to reduce the efficiency of anti-CD20 mAbs in targeting CNS resident B cells. Even when directly administered into the CSF (intrathecal), anti-CD20 mAbs have limited impact [13–15]. Because CNS resident B cells could play an essential role in disease progression, deeper CNS depletion of these cells might achieve greater efficacy relative to peripherally acting anti-CD20 therapies.

Chimeric Antigen Receptor (CAR)-T cells are genetically engineered cells approved for treatment of B cell malignancies and have a greater capacity for deeper tissue depletion of malignant B cells than antibody-based chemotherapy [16]. CD19-targeted CAR-T cells deplete B cells not only in the peripheral immune system but also within the CNS [17]. Recent anecdotal applications of CAR-T cells for several autoimmune diseases refractory to standard therapies suggest that deeper tissue depletion of B cells may have broad application in autoimmunity. Early experience in myasthenia gravis (MG) [18, 19], stiff-person syndrome [20], neuromyelitis optica spectrum disorder [21], and MS [22, 23] suggest that CAR-T cell treatments have acceptable risks of toxicity in neurologic conditions.

This study characterized the preliminary safety of mivocabtagene autoleucel (miv-cel, formally KYV-101), a fully human, autologous, CD19-targeting CAR T-cell therapy with CD28 co-stimulation [24]. This was studied in patients with progressive MS based on frequency and severity of adverse events (AE). Additional goals were a) to document CAR-T penetration into the CNS by confirming the presence of CAR-T cells in CSF following their peak expansion in peripheral blood, and b) to characterize changes in intrathecal gammaglobulin synthesis, a hallmark of abnormal humoral immune activation in MS. We also sought to understand disease activation and immunologic changes in the context of this type of deep B cell depletion using advanced molecular and cellular techniques such as single cell analysis, high-throughput discovery proteomics, viral seropositivity assays, and phage immunoprecipitation sequencing (PhIP-Seq). We combined these technologies to characterize broad changes in the immunologic landscape of MS and to investigate the effects of CAR-T cell therapy on compartmentalized inflammation and anti-Epstein-Barr Virus (EBV) immune response, give the association of EBV with MS pathogensis [25–28] and with compartmentalized inflammation [29].

## RESULTS

The study was designed to enroll ten participants to evaluate the penetration, safety and tolerability of a single dose of miv-cel. Kyverna Therapeutics stopped further recruitment after 2 participants were enrolled due to their shift in disease focus.

### Clinical course and adverse events

#### Participant 1

Participant 1 was a woman in her 50’s who was diagnosed in 2003 with RRMS and was untreated until 2017 when ocrelizumab was started. SPMS was diagnosed in 2022 based on insidiously declining ambulatory function. The expanded disability status scale (EDSS) score in 2023 was 3.5 and increased to 6.0 in 2024 (Figure 1B) despite ongoing ocrelizumab treatment and without clear evidence of relapse on MRI.

**Figure 1:**
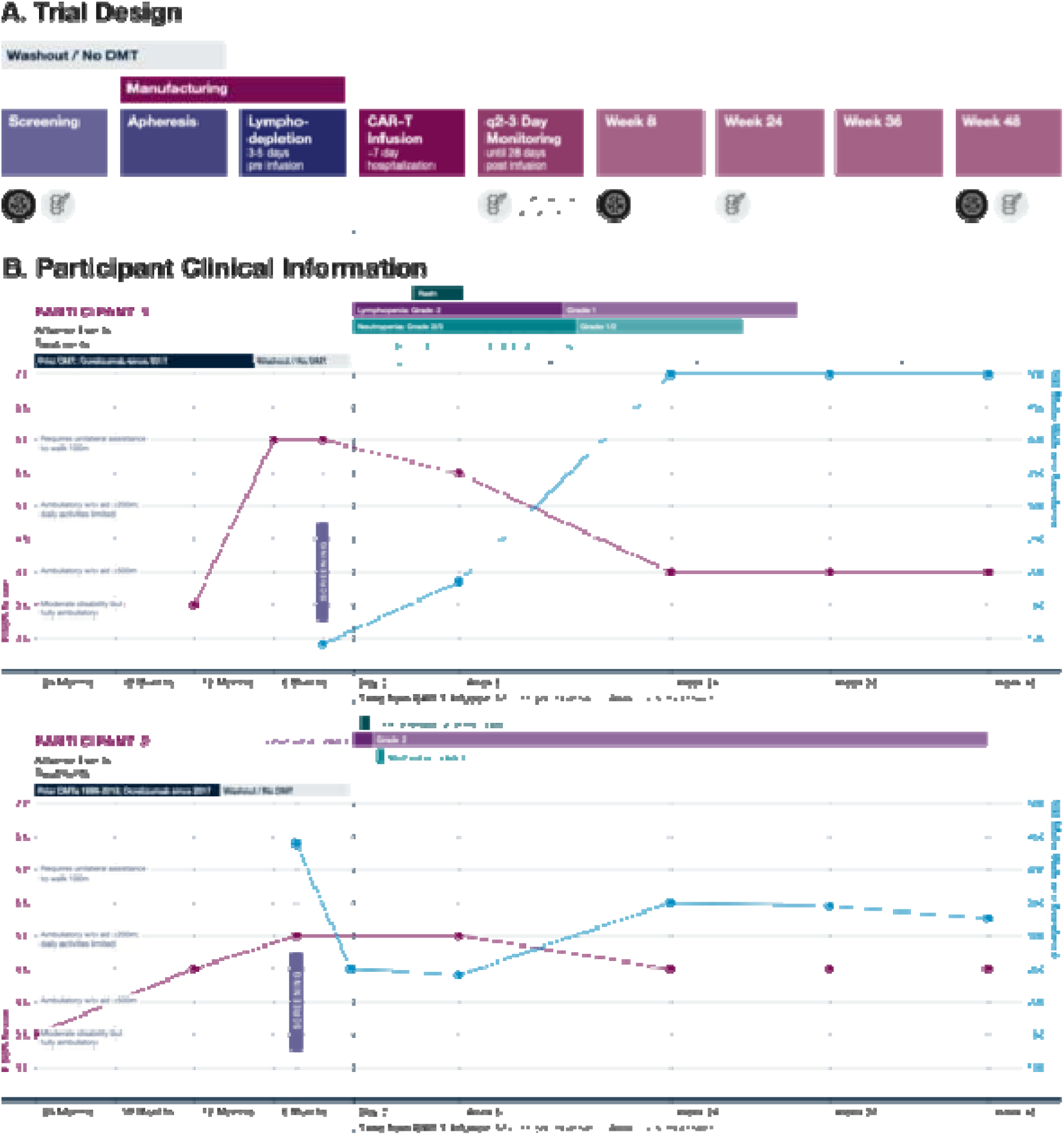
Trial Design and Participant Enrollment: **A.** Trial Design Schematic. Black circle with brain symbol denotes MRI. Syringe with vertebrae symbol denotes CSF collection **B**. Left: Participant’s EDSS changes leading to the diagnosis of treatment refractory multiple sclerosis. Right: clinical course after CAR-T including EDSS, 500m walk, and any AE.

Participant 1 was treated with 0.33 × 10^8 CAR+ T-cells at timepoint Day 0 after lymphodepleting chemotherapy on Day −5, −4, and −3 (fludarabine/cyclophosphamide). By Week 8, the participant’s EDSS decreased from 6.0 to 5.5 and then decreased further to 4.0 by Week 24 where it remained until end of study (EoS) at Week 48. The maximum distance ambulated increased from 90 meters with a cane at Screening to 185 meters with a cane at Week 8 to >500 meters by Week 24 when the participant began to walk unassisted. The timed 25-foot walk (T25FW) was 9.1 seconds on average from Screening to Week 8 and decreased to 5.9 seconds on average for Weeks 24-48 (Supplemental Figure 1A). There was no change in the paced auditory serial addition test (PASAT) or 9-hole peg test (9-HPT) (Supplemental Figure 1A). Neuroaxis MRI imaging and patient reported outcomes (PROs) for bowel/bladder and fatigue were unchanged from Screening to EoS (Supplemental Figure 1B).

Participant 1 did not experience cytokine release syndrome (CRS) or immune effector cell-associated neurotoxicity syndrome (ICANS) during the 7-day hospitalization. Starting Day 32 she developed progressive facial swelling, headache, and papular rash on the neck and chest; labs on Day 38 were notable for grade 3 transaminitis and a rapid rise in absolute lymphocyte count (ALC) from 0.26 x 10^9^/L on Day 32 to 1.09 x 10^9^/L. Dermatologic evaluation noted diffuse pruritic papules and plaques throughout in a descending pattern. Skin biopsy showed sparce superficial perivascular lymphocytic infiltrates that focally obscured the dermo-epidermal junction, where slight vacuolar change was present (Supplemental Figure 2A). Lymphocytes were CD3+ T cells with a mix of CD4 and CD8 positive cells (B cells were not identified). CAR-T cells were not detected based on droplet digital PCR (ddPCR) (Figure 2A). She was diagnosed with atypical CAR-related toxicity related to non-CAR T cell rise (further details found in Supplemental Table 1, Supplemental Figure 2B-C). In addition to topical hydrocortisone and barrier creams, she was treated with oral prednisone 40mg/day starting on Day 42 and tapered slowly over 4 weeks. The rash resolved by Day 59.

**Figure 2:**
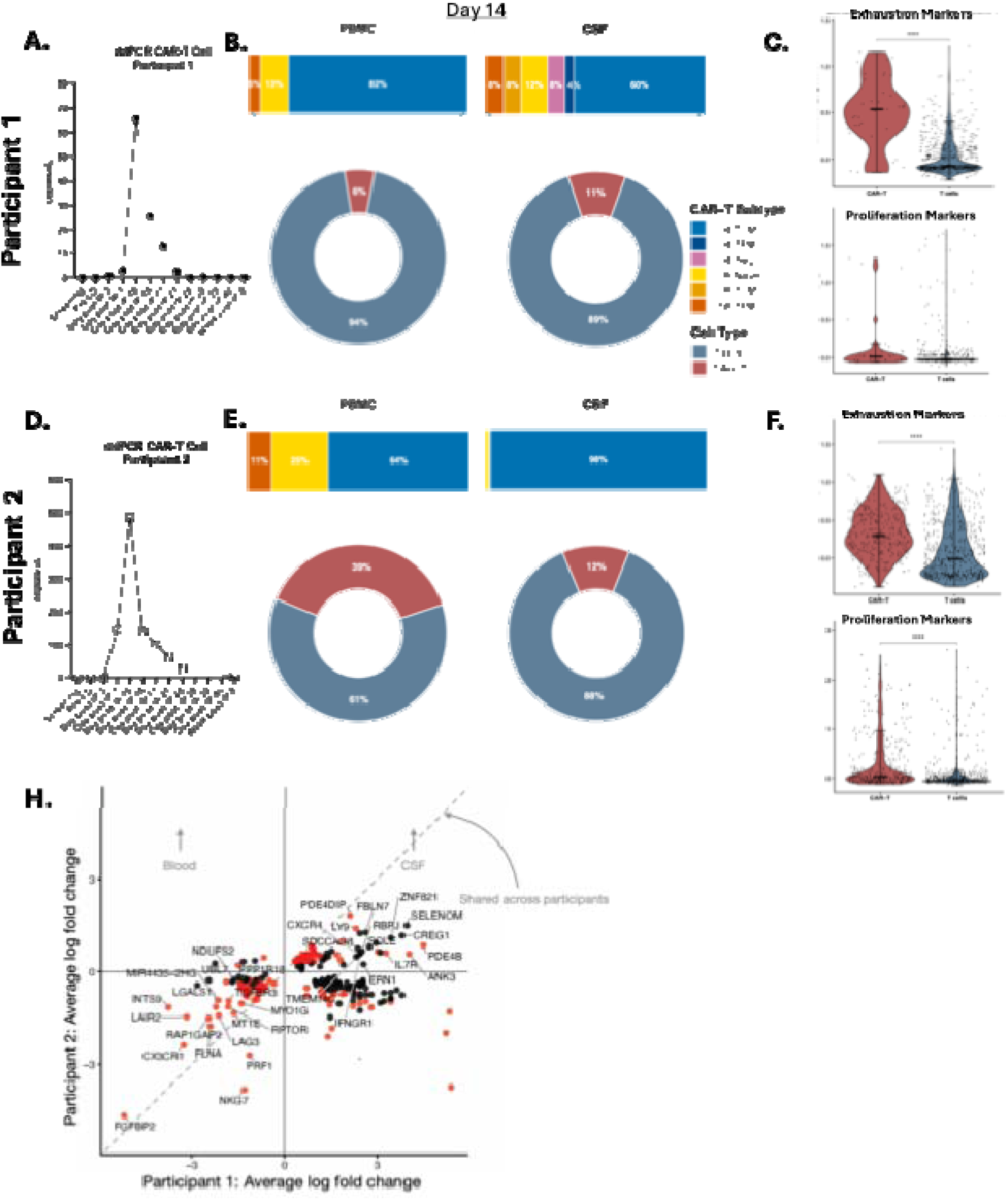
CAR-T Cell Dynamics: **A.** Participant 1’s longitudinal CAR-T copies/uL from Screening to Week 8 determined by ddPCR. **B.** Participant 1’s Day 14 CAR-T proportion in PBMC and CSF via single cell analysis and T cell subtypes based on single cell gene expression of CAR+ cells. **C.** Single cell gene expression abundance for proliferation (MKI67, TOP2A, PCLAF, CENPF, TYMS, NUSAP1, ASPM, PTTG1, TPX2, RRM2, CLSPN, TK1) and exhaustion (TIGIT, DUSP2, PAC1, PDCD1, LAG3, HAVCR2, TOX) markers from Day 14 in CAR+ and non-CAR-T cells from the peripheral blood. **** p < 0.0001 **D-F.** Same as A-C except for Participant 2. **H.** Differential Gene Expression of CAR+ cells in the CSF versus the PBMC across both participants (Participant 1 on the x-axis, Participant 2 on y-axis). Genes in the top right are expressed more in CSF CAR+ cells in both participants, genes in the bottom left are expressed more in the PBMCs, with the diagonal showing concordant genes. Red dots denotes p<0.05.

Participant 1 also developed grade 3 neutropenia (Day −1) requiring 7 doses of granulocyte-colony stimulating factor (G-CSF, filgrastim-sndz) for absolute neutrophil count (ANC) < 1 x 10^9^/L over four months (Days 21, 37, 70, 77, 83, 90, and 104), grade 3 lymphopenia (starting Day −1) and grade 2 hypogammaglobinaemia (starting at 94 days) requiring 0.5g/kg intravenous immunoglobulin (IVIg) 3 times (Days 101, 178, 287) for IgG levels < 400mg/dL, and chemotherapy-induced alopecia. The neutropenia, lymphopenia, and alopecia resolved 6 months post CAR-T infusion (Figure 1B).

#### Participant 2

Participant 2 was a woman in her 60’s diagnosed with RRMS in 1997. She was treated with interferon beta-1 from 1999-2013 and then dimethyl fumarate from 2013-2015. Due to persistent lymphopenia on dimethyl fumarate, rituximab was started in 2016, and subsequently transitioned to ocrelizumab in 2017. SPMS was diagnosed in 2018 due to gait decline. Participant 2’s EDSS score was 3.5 in 2023 and increased to 4.5 in 2024 despite ongoing treatment without acute MRI changes. While being off all disease modifying therapy (DMT) for nearly one year during washout and while awaiting CAR-T manufacturing delays, her EDSS score increased to 5.0 immediately pre-CAR-T (Figure 1B).

Participant 2 was treated with 0.33 × 10^8 CAR+ T-cells at timepoint Day 0 after the same lymphodepleting treatment as participant 1. Improvements in function were noted, and the EDSS decreased to 4.5 by Week 24 and remained stable until EoS (Figure 1B). Maximum distance ambulated at Screening was 440 meters, which declined to 250 meters following CAR-T infusion and then improved to 320-350 meters unassisted by Week 24 through EoS. The T25FW, PASAT, 9-HPT, neuroaxis MRI imaging and PROs were stable from Screening through EoS (Supplemental Figure 1A-B).

She experienced grade 1 CRS on Day 4 presenting with fever and concurrent pseudo-relapse of prior MS symptoms including right leg weakness and bilateral leg paresthesias. She was treated with acetaminophen followed by tocilizumab on Day 5 due to ongoing fever, and her symptoms resolved. Recurrent fever on Day 6 and was treated with dexamethasone 10 mg and defervesced. On Day 8, fever recurred and tocilizumab was administered with resolution of her fever. She was discharged on Day 10. She subsequently experienced grade 4 lymphopenia for 10 days and maintained a grade 2 lymphopenia until EoS. Post-discharge, she endorsed worsened fatigue; however, two months after treatment and with initiation of physiotherapy, she noted improvements in fatigue and mobility to pre-enrollment levels.

### CAR-T Cell Dynamics

The CAR-T infusion product for participant 1 was 62% CAR+ and was made up of 54.7%:41.0% CD4:CD8 CAR+ T cells. Participant 2’s product was 56% CAR+ and made up of 83.0%:15.6% CD4:CD8 CAR+ T cells. The breakdowns of the CAR+ T cell subtypes for participants 1 and 2 were 30.8% and 40.4% central memory CD4, 68.3% and 59.5% effector memory CD4; 25.8% and 49.5% central memory CD8, 69.1% and 48.9% effector memory CD8, respectively.

Using ddPCR of whole blood to measure copies of CAR construct per microliter during the first 8 weeks, CAR-T expansion for participant 1 was detectable on Day 3, peaked on Day 10 and became undetectable by Day 28 (Figure 2A). For participant 2, CAR-T expansion was detectable after Day 3, peaked on Day 10, and became undetectable by Week 8 (Figure 2D).

Based on CAR detection by RNA-Seq/CITE-Seq at Day 14, the CAR-T cells represented 6% of all T cells in peripheral blood and 11% in CSF for participant 1 (Figure 2B) and 39% and 12%, respectively, for participant 2 (Figure 2E); peripheral blood CAR+ CD4:CD8 percentages changed from the infused product to 82%:18% for participant 1 (from 54.7%:41.0%) and 64%:36% in participant 2 (from 83.0%:15.6%). The CAR-T cell subtypes for participant 1 in peripheral blood were CD4 central memory (82%), CD4 effector memory (5%), and CD8 naïve (13%) (Figure 2B, Supplemental Figure 3E). The peripheral blood CAR-T cell subtypes in participant 2 were CD4 central memory (64%), CD8 naïve (25%), and CD8 effector memory (11%) (Figure 2E, Supplemental Figure 3E). In the CSF, there was a CD4 predominance for both participants: 72% for participant 1 and 98% for participant 2. In participant 1, the CSF CAR-T composition was CD4 central memory (60%), CD4 effector memory (4%), CD4 T regulatory cells (8%), CD8 naïve (12%), CD8 central memory (8%), and CD8 effector memory (8%) (Figure 2B). In participant 2, the subset proportions were skewed towards CD4 central memory (98%), with minimal CD4 effector memory (0.2%), and CD8 naïve (1.8%) (Figure 2E). No clonal expansion was noted in the CAR+ populations on Day 14 as assessed by single cell T cell receptor (TCR) sequencing in both participants in both peripheral blood and CSF (Supplemental Figure 3C-D).

On Day 14, the peripheral blood non-CAR-T cells in both participants had similar subtype breakdowns of the CAR+ cells except for the ongoing presence of (non-CAR-T) CD4 T regulatory cells (8% in participant 1 and 5% in participant 2). The CSF CAR-T and non-CAR-T cells subtype breakdown was similar (Supplemental Figure 3A-B).

Proliferation and exhaustion markers had higher expression levels in CAR-T cells compared to the non-CAR cells in peripheral blood (Figure 2C, 2F, p <0.001) except for participant 1’s proliferation gene expression program which showed a trend of upregulation that was not statistically significant (p = 0.0647). As expected, T cell trafficking markers had a higher expression level in T cells in the CSF compared to PBMCs and were overall unchanged from Screening (∼20-40% vs 0-10% of T cells expressing trafficking markers, respectively) (Supplemental Figure 3F-G). Differential gene expression (DGE) analysis of CAR-T cells in the peripheral blood versus CSF at Day 14 revealed increased expression of genes associated with T cell activation and cytotoxicity were upregulated in peripheral blood when compared to CSF (PRF1, FGFBP2, CX3CR1, NKG7) (Figure 2H). In CAR-T cells from the CSF relative to peripheral blood,, there was a higher expression of genes related to trafficking and memory-survival (CXCR4, IL7R, IFNGR1) (Figure 2H). Interestingly, in a Reactome pathway analysis of CAR-T cells in each compartment, viral translation genes were upregulated in the CSF, potentially in the context of virally integrated CAR expression (Supplemental Figure 3H). DGE differences between CSF CAR-T compared to CSF non-CAR-T cells showed higher expression signatures of receptor activation (LYN, GAB2), proliferation (AURKA), and exhaustion marker (NR4A2) in CAR-T cells suggestive of local antigen engagement (Supplemental Figure 3I).

No CAR-T cells were detected by ddPCR past Week 8; however, there were minimal CAR-T cells noted at Week 48 only by scRNA-Seq (<1% of total T cells). In participant 1, 3 CD4 naïve, 8 CD4 T central memory, 2 CD8 naïve, and 1 CD8 T central memory CAR+ cells were found in the peripheral blood and 3 CD4 T central memory and 1 CD8 naïve CAR+ cells in the CSF. In participant 2, 3 CD4 T central memory CAR+ T cells in the peripheral blood and 1 CD4 T central memory in the CSF were found. None of the 18 CAR-T cells identified across both participants could be tracked back to the initial expansion on Day 14 based on TCR repertoire analysis, suggesting unrelated clonotypes. Though this is limited data, this is different from the oligoclonal CAR persistence described in adult oncology [30–32].

Overall, the expanded CAR+ cells in the peripheral blood were similar to the subtypes in the infused product and a representation of the non-CAR population except for T regulatory cells. CAR+ cells in the CSF had a CD4 predominance with increased trafficking markers. There was evidence of overall CAR-T cell activation and target engagement in both compartments.

### B Cell Reconstitution

Both participants were treated for over five years with anti-CD20 monoclonal antibodies prior to trial entry. At baseline, CD19+ B cells were not detected in the peripheral blood by clinical lymphocyte subsets, flow cytometry, or scRNA-Seq. Around Week 12, both participants began to have detectable B cells such that by Week 24, both participants’ peripheral B cells were within normal limits (Figure 3A). In the CSF, by scRNA-Seq at Weeks 24 and 48, only participant 1 had B cells detected at low numbers of 15 and 40 cells, respectively (Figure 3A).

**Figure 3:**
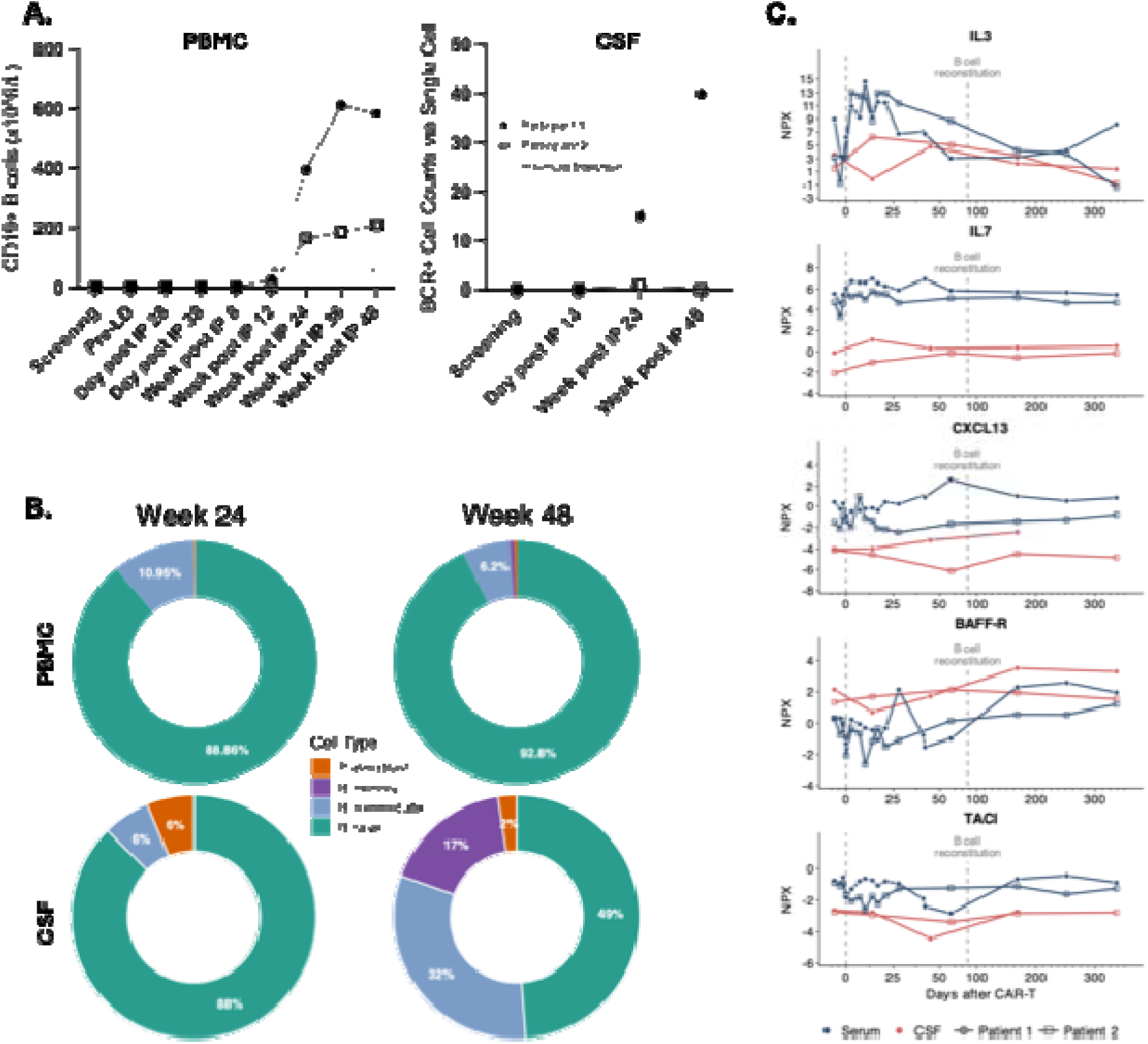
B cell Dynamics: **A.** (Left) Longitudinal plot of CD19+ cells from peripheral blood based on CLIA Certified Lymphocyte Subset Assay. (Right) Longitudinal plot of of BCR+ counts based on single cell in the CSF. **B.** B cell subtypes from Weeks 24 and 48 based on single cell gene expression, combined for participant 1 and 2 **C.** Longitudinal normalized protein expression (NPX) levels of CXCL13, B-cell activating factor receptor (BAFF-R), and transmembrane activator and CAML interactor (TACI), measured by proximity extension assays, in serum (blue) and CSF (red) from participant 1 (filled circle) and participant 2 (open square). For clarity, the early post-CAR-T period (Day +1 to +56) is expanded approximately threefold on the x-axis relative to the pre-CAR-T and late post-CAR-T periods. The vertical dashed line at Day +84 indicates the time of B-cell reconstitution. For CXCL13 and TACI, one CSF data point from participant 1 was excluded following assay-specific quality control.

The reconstituting peripheral B cells were primarily naïve with 60-70% having the cell surface marker pattern CD45+CD3-CD19+CD24hiCD38hi based on flow cytometry at Week 24 (Supplemental Figure 4A). In both participants, scRNA-Seq confirmed a naïve reconstitution at Week 24 with 88% naïve with an additional 6-10% intermediate subtype in both the periphery and CSF (Figure 3B). By Week 48, there were relatively higher proportions of intermediate and memory B cells in the CSF compared to CSF at Week 24 and to peripheral blood at Week 48 (Figure 3B-C). Single cell B cell receptor (BCR) analysis provided additional evidence for the ongoing reconstitution of naïve B cells from Weeks 24 to 48 with an absence of antigen specific B cell maturation and no persistence of distinct B cell clones (Supplemental Figure 4B).

At Week 24, both participants at Week 24 showed significant reconstitution in peripheral B cells of mainly IgM isotype and very few CSF B cells (12 IgM and 1 IgD naïve in participant 1 and 4 identical IgG1 with 11.5% VH mutation in participant 2). At week 48, peripheral and CSF B cells in participant 1 had further reconstituted and were predominantly IgM+, whereas participant 2 exhibited largely unchanged peripheral B cell levels and no detectable CSF B cells. There was no bias in heavy or light chain germline family usage or CDR3 length changes during treatment (Supplemental Figure 4C-E) and no common heavy chain variable domain sequences were found between timepoints or participants.

In the CSF of participant 1 there were no clonal families (CF) at Screening or up to Week 24. At Week 48, 1 IgM CF (n=6, no mutations) and 1 IgA1 CF (n=2, 2% mutation) were measured. Participant 2 had one IgG1 CF of size of 4 with 11.5% mutation at Week 24 and no B cells at Week 48. In the blood clonal families were not present in either participant from Screening up to Week 8. At Week 24 participant 1 had 53 CF (2-5; 0-2%; 0-2% mutation) and 83 CF (2-33; 0-7% mutation) at Week 48, mainly IgD/IgM, IgM and IgA1. Participant 2 had 18 CF (2-5; 0-2.6% mutation) at Week 24, mainly IgM, 1 IgD/IgM and 2 IgA and 15 CF (2-6; 0-2.7% mutation) at Week 48 of 1 IgD, 7 IgM and 7 IgA1 isotypes. There was no evidence of clonal family molecular evolution/antigen maturation in larger CFs, members showed an expansion of a single clone (Supplemental Figure 4E)).

### B Cell Homing

To identify drivers of naïve B cell reconstitution, proteomics and gene-expression markers were longitudinally tracked, including markers of bone marrow activation and primitive cell differentiation (IL-3, MME, and CXCR4); B cell lineage commitment and VDJ rearrangements (IL-7, PAX5, TCL1A, CD24, and CD38); bone marrow egress (S1PR1, SELL); lymphoid homing (CXCL13, CXCR5, MS4A1, FCRL1, FCER2); B cell maturation and survival (BAFF, BAFF-R (TNFRSF13C), 4-1BB, TACI (TNFRSF13B), B-cell maturation antigen (BCMA, TNFRSF17)) [33–39].

When looking at longitudinal protein analysis, for both participants exhibited a significant rise in serum IL-3 and IL-7 “early” after CAR-T infusion (Figure 3C, Supplemental Figure 6H). For participant 1, serum CXCL13 levels began increasing at Day 28, prior to the detection of peripheral B cells. BAFF-R levels peaked transiently around Day 28, shortly after participant 1 received the first GM-CSF dose, and subsequently increased gradually to above-baseline levels in serum and CSF, coinciding with peripheral B cell reconstitution (Figure 3C). In participant 2, CXCL13 transiently peaked within the first 10 days after CAR-T infusion and then showed a gradual increase beginning after Day 28. BAFF-R increased in the serum, but not CSF consistent with her B cell reconstitution. In both participants, emergence of B cells corresponded to a slight decline in serum BAFF levels and a marked increase in serum CD22, FCER2, and FCRL1 levels (Supplemental Figure 5A), consistent with B cell reconstitution. TACI and 4-1BB exhibited a steady state after B cell reconstitution, both in serum and CSF (Figure 3C, Supplemental Figure 5A).

DGE analysis focusing on markers of B cell maturation and reconstitution (PAX5, SELL, MS4A1, MME, CD38, CD24, and TCL1A), and B cell survival and homing (CXCR4, S1PR1, CXCR5, BCMA (TNFRSF17), TACI (TNFRSF13B), BAFF-R (TNFRSF13C) were explored. Peripheral blood showed no changes from Week 24 to 48, matching the ongoing >90% representation of a naïve proportion of B cells at Week 48 (Figure 3B) and a new BCR repertoire (Supplemental Figure 5B). There was a relative rise from Week 24 to 48 in both maturing and survival markers in the CSF (Supplemental Figure 5B), mirroring the later differentiated B cell subset in the CSF at Week 48 (Figure 3B). In addition, there was a rise in expression levels of S1PR1, higher levels of CXCR4 and higher levels of BAFF-R (Supplemental Figure 5B).

### Immunologic Impact Post-CAR-T

#### T cells

In both participants in the peripheral blood, based on scRNA-Seq, non-CAR+ T cells decreased by Day 14 with loss of CD4 and CD8 naive cells (Figure 4A, Supplemental Figure 6A), consistent with lymphodepletion effects [40] and CAR-T expansion. As numbers began to rise again, starting Week 8, there was an increased proportion of the CD4 T central memory and CD8 T effector memory populations (Figure 4A, Supplemental Figure 6C-D). By TCR repertoire analysis, there were nominal shared clones from Screening to Week 48; however, there were more shared clones between blood and CSF at Week 48 (Supplemental Figure 6B).

**Figure 4:**
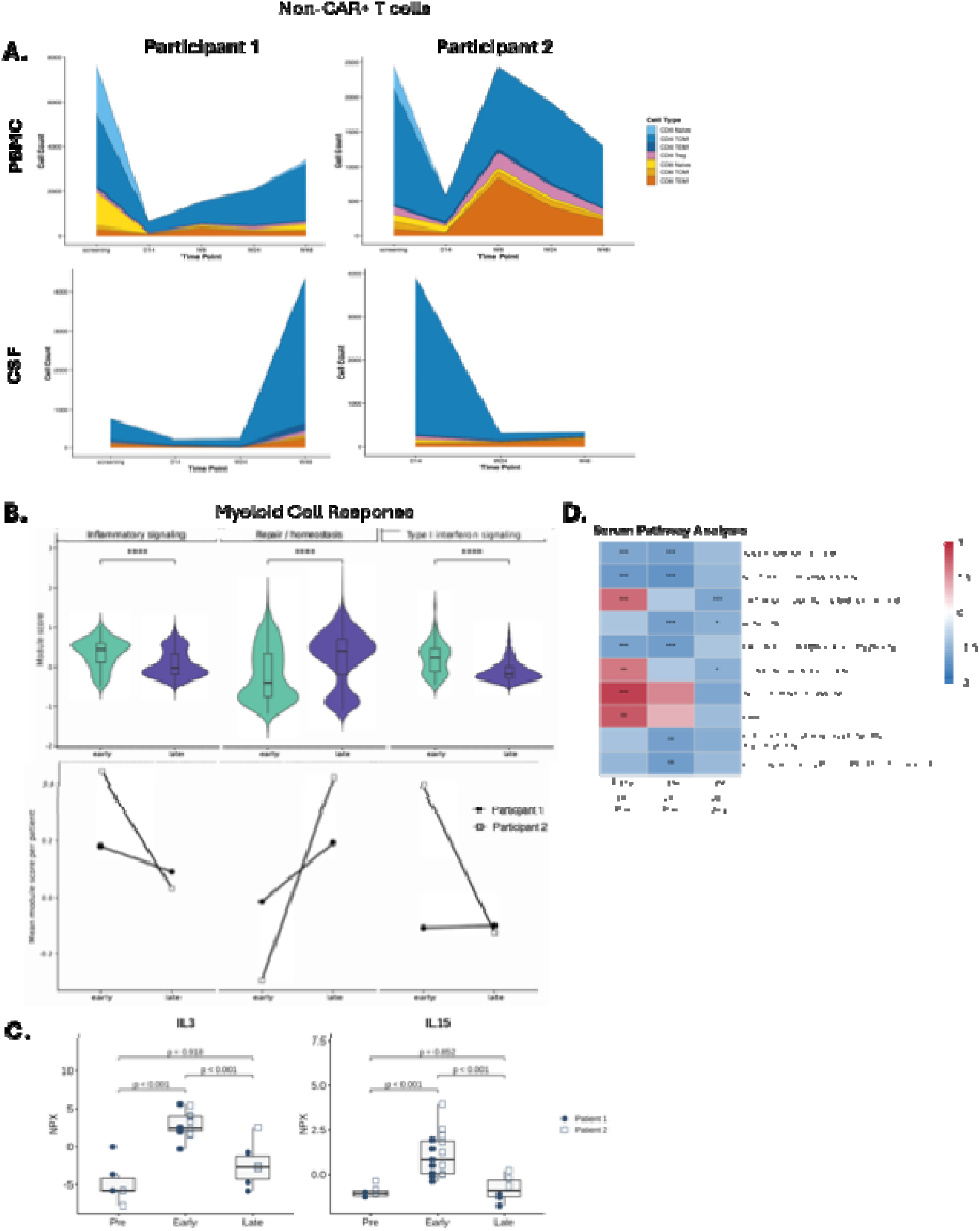
Post-Treatment Immunologic Changes: **A.** Based on non-CAR CD3+ T cells, relative abundance and subtypes longitudinally based on single cell gene expression. **B.** Based on single cell, isolating the myeloid compartment and looking at gene expression changes with a focus on activation state. **** p < 0.0001 **C**. Normalized protein expression (NPX) levels for IL-3 and IL-15, measured by proximity extension assays, during the pre-CAR-T, early post-CAR-T (Day +1 to +56), and late post-CAR-T (> Day +56) period in participant 1 (filled circle) and participant 2 (open square). Boxplots show the median (center line), interquartile range (box), and values within 1.5 × IQR (whiskers). Nominal p-values were derived from *limma* analysis. **D.** Heatmap displaying proteomic pathway enrichment across the pre-CAR-T, early post-CAR-T, and late post-CAR-T periods. Shown are the normalized enrichment scores (NES) of the 10 most consistently enriched pathways from gene set enrichment analysis (GSEA) using the Reactome collection across three pairwise comparisons (Early vs. Pre-CAR-T, Late vs. Pre-CAR-T, Late vs. Early). Pathways were ranked by the number of statistically significant comparisons (p_adj_ < 0.05) and, within each rank, by the smallest adjusted P value. Red and blue indicate positive and negative enrichment, respectively, relative to the reference group. For example, a blue color for “Early vs. Pre-CAR-T” indicates lower pathway enrichment during the early post-CAR-T period than during the pre-CAR-T period. * p_adj_ < 0.05; ** p_adj_ < 0.01; *** p_adj_< 0.001.

For participant 1, T follicular helper (Tfh) gene cell markers (CXCR5, PDCD1, ICOS, BCL6, MAF, CXCL13) had stable expression in both blood and CSF from Screening to Week 48 (Supplemental Figure 6E). Participant 2 showed a reduction in expression levels of Tfh cells in her blood (Supplemental Figure 6E). Participant 2’s CSF Tfh gene expression levels were similar to participant 1’s CSF expression levels at both time points despite their clearance of oligoclonal bands or normalization of IgG index.

There was no consistent increase or decrease in virus-associated TCR frequency before versus after CAR-T therapy (Day 14-Week 48) in either participants or compartments (peripheral blood vs CSF) when aligning TCR sequences to cytomegalovirus (CMV), Influenza, or SARS-CoV-2 from VDJdb datasets (Supplemental Figure 6F). EBV-aligned TCRs were undetectable in both participants, which may have been influenced by prior anti-CD20 treatment. These findings did not change after a rise in T cells or B cell reconstitution (Supplemental Figure 6F).

#### Other Immune Cells

Analysis of DGE between early (Screening and Day 14) and late timepoints (Weeks 24 and 48) in the CSF myeloid compartment showed significant upregulation of translation pathways in the late timepoint and significant downregulation of interferon signaling pathways, toll like receptor 4 pathway and the DAG IP3 pathways, consistent with a less inflammatory state (Figure 4B, p <0.0001).

Following the CAR-T cell expansion phase, serum proteomics showed that proinflammatory markers that had peaked early after CAR-T cell infusion, including IL-3 and IL-15, declined to baseline levels (Figure 4C, Supplemental Figure 6H). At the pathway level, an even broader and more sustained attenuation of inflammatory responses was observed, with pathways related to innate immune activation and neutrophil degranulation remaining downregulated at late timepoints (>Day 56) compared with pretreatment (Figure 4D). While there was treatment associated neutropenia in participant 1, this downregulation was still noted at the “late” stage when her neutrophils were essentially normal. Similarly, IL-10 signaling remained upregulated throughout the late post-CAR-T phase, although this did not retain significance after FDR correction (Figure 4D). In the CSF, participant 2 exhibited a gradual decline in IL17A, IFN-γ, MMP9, GM-CSF and IL2RA (Supplemental Figure 6I) after treatment, supporting an overall attenuation of CNS inflammation, which was not consistently observed for participant 1. Markers of CNS injury (GFAP, NFL) and microglial activation (SPP1) were stable in both CSF and serum during the trial duration (Supplemental Figure 6I).

### CD19 Target Engagement and Antibody Changes

Both participants had unique OCBs within their CSF at the time of Screening. In the CSF of participant 1, 1 unique and 3 matched bands were seen at Screening which were later reported as 4 unique OCBs on Day 14. After treatment, participant 1 had no unique OCBs at Weeks 24 or 48 (Figure 5A), and CSF kappa free light chain was undetectable by Week 48. The IgG index was normal and unchanged (Figure 5A). Participant 2 had >5 unique OCBs at Screening (Figure 5A). Neither OCBs nor kappa free light chain were affected by CAR-T infusion (Figure 5A). However, the IgG index decreased from 0.71 at Screening to 0.57 at Week 48 (Figure 5A). Serum vaccine titers declined for participant 1 but were unchanged in participant 2 (Supplemental Figure 7).

**Figure 5:**
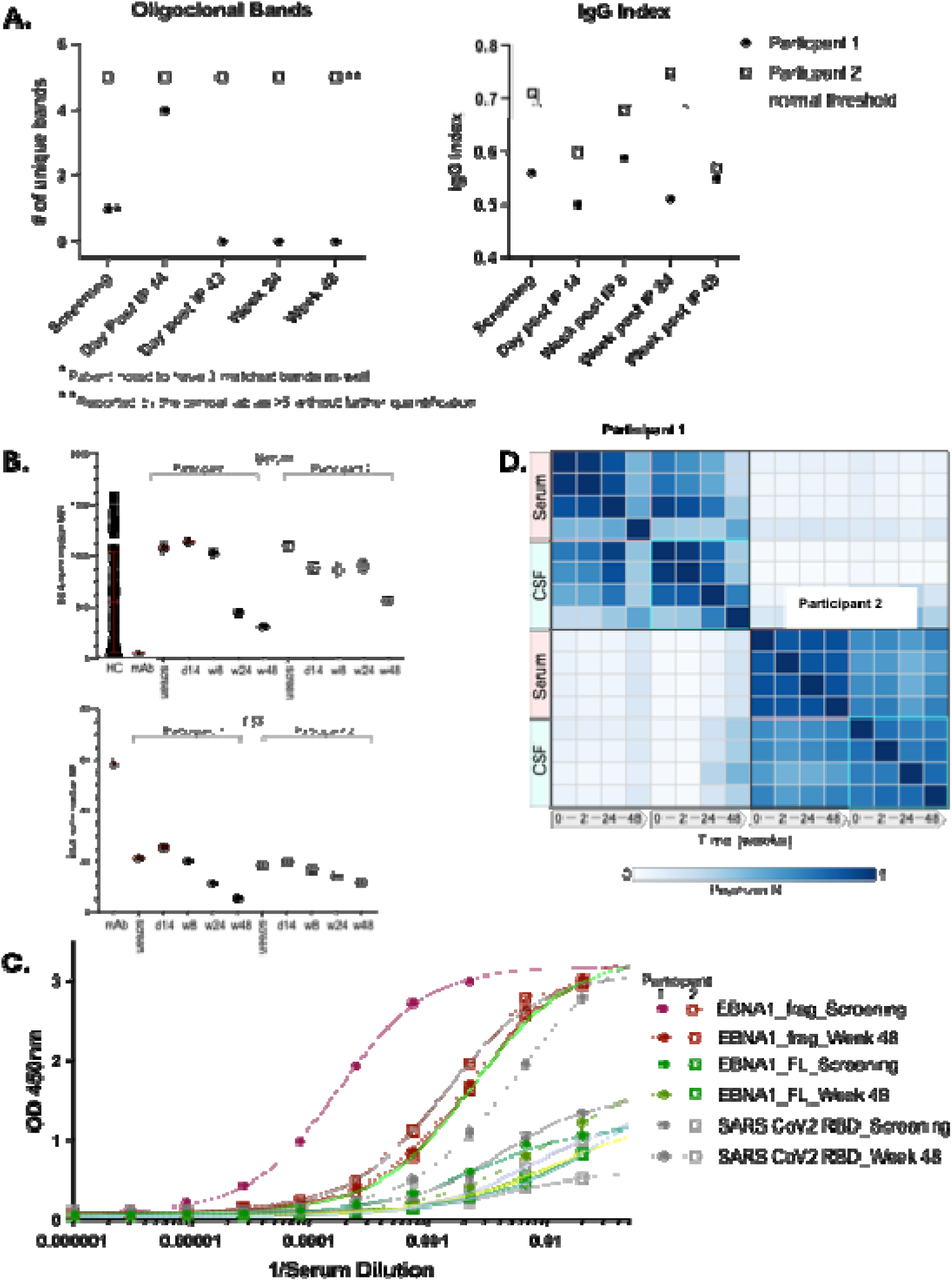
CD19 Targeting and Antibody Outcomes. **A.** Longitudinal CLIA Certified oligoclonal band and IgG index assay results based on serum and CSF results. **B.** IgG directed to a BSA-peptide conjugate of EBV EBNA1 amino acids 393-412 was quantitatively measured as BSA normalized mean fluorescence intensity (MFI) using Luminex. Serum (upper plot) diluted 1/250 or CSF (lower plot) diluted 1/40 were compared over the treatment course for each participant and to sera from healthy controls. A monoclonal antibody to the peptide was used as a control in the assay at 0.5 ng/mL. **C.** IgG serum titers to a GST-EBNA1 fusion protein (red; amino acids 386-500), full length EBNA1 (green) or SARS CoV2 RBD (grey) were measured by ELISA for each participant at the Screening and Week 48 timepoints. **D.** Phage ImmunoPrecipitation Sequencing Autoreactome Pearson R correlation of longitudinal serum and CSF.

#### Epstein-Barr Virus (EBV)-Associated B Cell Changes

EBV serologic reactivity before and after CAR-T treatment was examined. Both participants were seropositive to an EBV EBNA1 peptide (amino acids 393-420, i.e., EBNA1.frag1) associated with MS risk [41–46]. In serum, participant 1 showed a 46% drop in seroreactivity from baseline to Week 48. Participant 2 had a 20% drop in seroreactivity (Figure 5B). Both participants were also seroreactive in CSF to the EBV EBNA1.frag1 peptide [41–46]. Participant 1 showed an 82% drop in signal by Week 48 whereas participant 2 seroreactivity decreased by 20% (Figure 5B). When looking at the anti-EBNA1 C-terminal fragment (amino acids 386-500) antibody titers, participant 1 had a >10-fold drop in titers at Week 48 compared to Screening (Figure 5C) despite a rise in other antibodies such as SARS-CoV-2 (post revaccination). Participant 2 EBNA1 fragment antibody titers did not change significantly.

#### Other antibody-based trends

The human proteome-wide phage immunoprecipitation sequencing (PhIP-Seq) autoreactivity profiles [47], looking at the naturally occurring self-reactive antibodies in the serum demonstrated no major change in blood, as previously found with anti-CD19 CAR-T treatment [47], or in CSF over the course of the study except for a fluctuation at Week 48 for participant 1 which could not be ruled out as secondary to IVIg treatment (Figure 5D).

## DISCUSSION

This early CAR-T study followed two treatment refractory progressive MS participants post anti-CD19 CAR-T for over a year. The primary endpoint was met: CAR-T cells were detected in the CSF following peak expansion. The secondary endpoint of characterizing target engagement based on normalizing intrathecal gammglobulin synthesis was also met with clearance of OCB for participant 1 and normalization of the CSF IgG index for participant 2. We found central memory CD4 predominant CAR-T expansion in the CSF, peripheral naive B cell reconstitution starting at Weeks 8-12, and a less inflammatory background by Week 48 compared to Screening.

To date, two other publications have reported CAR-T cell therapy in multiple sclerosis. Fischbach, et al. had an early interim report of two progressive MS participants treated with the same product, miv-cel (formally KYV-101), at a dose of 1×10^8 cells [48]. Both participants showed CAR-T expansion in the periphery and the CSF. The first participant developed grade 1 CRS associated with facial and neck swelling, and was found to have a new MRI lesion in the thoracic spine at day 64. This same participant had a reduction in OCB and improvement in long distance walking. The second participant develop no toxicity and had no change in the OCB, IgG index, exam, or MRI though follow-up time was limited. Qin, et al. published an interim report in five participants with MS treated with 1×10^6/kg of anti-BCMA CAR-T cells [23]. All participants showed CAR-T expansion, including prolonged expansion in the CSF with a CD4 predominance. Only grade 1 CRS was reported (in 4 of 5 participants), but all participants experienced grade ≥ 3 cytopenias. There was partial reduction in OCB and clearance of kappa free light chain, and clinical outcomes improved alongside evidence of CNS myeloid downregulation post treatment. The results here are overall similar to the findings reported in these two publications in terms of CAR-T expansion, toxicities, and clinical outcomes.

The CAR-T cells expanded well despite using one third of the typical dose used in oncology or other clinical trials for patients with autoimmune disorders [20–23, 49–51]. Expansion was mainly driven by CD4 central memory cells becoming the predominant phenotype in all compartments in both participants, with no clonal expansion of the CAR-T TCR repertoire and generally close maintenance of pre-existing T cell subtype proportions outside of T regulatory cells. Participant 2 had nearly 10-fold higher CAR-T expansion compared to participant 1, potentially due to greater numbers of CD4 cells present which are associated with increased proliferation [52] or perhaps due to a less differentiated population with a higher proportion of central memory cells [52]. Despite the expansion, the grade and duration of the hematologic AE were more prominent in participant 1 indicating that these events are not directly linked to CAR-T expansion but more likely due to idiosyncratic host responses.

The repertoire analysis supported that the CSF CAR-T cells were not a dominant peripheral clone or a monoclonal line that expanded within the compartment but rather represent trafficking into the CSF. DGE showed that CSF CAR-T cells had higher expression of trafficking markers compared to peripheral CAR-T cells. CSF CAR-T cells showed expression of markers consistent with target engagement when compared to non-CAR CSF T-cells. This is all suggestive of compartment specific expansion and engagement. These markers should be studied more in larger numbers to help understand CNS CAR-T penetration.

Mirroring CAR presence in the CSF, eradication of OCBs was observed in participant 1. This was consistent with other antibody changes such as hypogammaglobinemia and relative decrease in clinical vaccine titers. Although OCBs were not eradicated, intrathecal synthesis of gamma globulins normalized in participant 2 as measured by the IgG Index. The reasons for the different impact of CAR-T treatment on CSF restricted OCBs in the two participants are not immediately apparent. It is possible that plasma cells that no longer express CD19 produce OCBs in a subset of MS patients. It is also possible that a single low dose of anti-CD19 CAR-T cells may fail to eradicate resident B cells within the CNS responsible for antibody production only in some individuals. Tfh cells are also present in ectopic follicles and might contribute to CNS compartmentalized inflammation [53–55]. For both participants, CSF Tfh cell gene expression was unchanged by CAR-T treatment.

MS patients exhibit enhanced antibody production to a C-terminal region of EBNA1 [41–46, 56] that presumably reflects increased immunologic response potentially due to increased EBV levels. Antibody titers against this EBNA1 region decreased following CAR-T infusion in both participants in blood and CSF. EBV specific TCR representation remained low, presumably due to prior anti-CD20 monoclonal antibody treatment [57, 58]. While unclear as to the long-term benefit of reduced EBV burden in progressive MS [59], this could be another marker to follow for treatment response.

Typically, following several years of anti-CD20 monoclonal antibody treatment, peripheral B cell counts become detectable only after 12-18 months or even longer after treatment discontinuation [60–63]. However, following CAR-T treatment, complete reconstitution was achieved by Week 24 despite the participants having undetectable peripheral B cells for years before receiving CAR-T treatment. We hypothesize that bone marrow B cell maturation is first driven by the elevation of cytokines such as IL-3 (progenitor differentiation) and IL-7 (early B cell maturation) associated with CAR-T expansion (noted in cytokine assays and in proteomic data). The elevation of S1PR1 found at later time points is associated with a push for immature cells to egress the bone marrow. The elevation of CXCL13 in participant 1 and BAFF-R in both participants was likely acting as a lymphoid attractant directing B cells to lymph nodes and CXCR4 as a lymph node retainer. The delayed increase of TACI after Week 24 may be consistent with a maturing B cell population over time. Deeper depletion of tissue resident B cells might provide a greater stimulus to the bone marrow to produce B cells more than is possible by anti-CD20 monoclonal antibody treatment. Interestingly, the anticipated rise in serum BAFF levels (noted following anti-CD20 B cell depletion [64]) was not found after CAR-T cell treatment. This lack of rise may be due to prolonged anti-CD20 monoclonal antibody therapy prior to CAR-T treatment resulting in an already elevated state of BAFF [64, 65].

By Week 24, both participants showed improvement in EDSS and 500m walk from pre-treatment, and this improvement sustained until Week 48 despite not using other immunosuppressive treatment as well as the return of normal levels of B cells in the periphery. The potential of sustained benefit post CAR-T treatment was supported by Week 48 reduction in inflammatory milieu of both participants based on the myeloid cell markers and general proteomic pathway analysis. Longer follow up is needed to assess the potential for immune reset and long-term remission.

Although participant 1 did not experience CRS or ICANS, an AE of transaminitis, facial swelling, and a systemic rash occurred. The CAR-T cell counts, the different TCR clonal expansion, proteomic analysis (found in supplemental table), and T cells within the dermatologic biopsy support an endogenous T cell rebound effect after CAR-T cell contraction. Despite extensive investigation, no insight was gained into why this AE occurred. It is possible this was a delayed drug reaction to either CAR-T or supportive management such as G-CSF versus an unmasked autoimmune process in a patient with known autoimmunity. This dermatitis was inconsistent with the recently described local immune effector cell-associated toxicity syndrome (LICATS,[66]) due to lack of pathologic B cells in the tissue effected. Non-CAR+ cells that were part of the infused product (38-44% of the T cells infused) were unlikely to cause dermatitis because scRNA-Seq showed an increase in the diversity of TCR repertoires at Day 14 and a change in the subtype represented in the non-CAR+ group compared to Day 14. Overall, this was suggestive of an alternative type of AE that should continue to be considered as more participants are treated with CAR-T cells in autoimmunity. In addition, in both participants, manageable yet prolonged cytopenias were observed. This may be due to prior prolonged exposure to anti-CD20 monoclonal antibody therapy which has been associated with bone marrow fatigue post CAR-T resulting in protracted cytopenias in the oncologic experience [67] and may be an important consideration in MS patients receiving CAR-T.

CAR-T cells are an exciting avenue of therapeutic development in the treatment of autoimmunity including MS. The limited yet varied experience in treating two participants highlights the importance of deeply characterizing the immunologic processes occurring in CAR-T treated MS participants to better understand what drives CAR-T expansion, penetration, and clearance, and how these processes influence therapeutic outcomes, toxicities, and sustainability of benefit.

## MATERIALS AND METHODS

Study Design and Oversight description is found in Supplemental Methods. The full study protocol is also found among the supplemental materials.

### Single Cell Analysis

Single cell sequencing sample processing is outlined in Supplemental Methods. All raw FASTQ sequencing data were processed using Cellranger (v9.0.0, multi). This included scRNA-Seq, scTCR-Seq, scBCR-Seq and Cell Surface Protein (CSP) data. Human reference transcriptome *GRCh38-2020-A*, V(D)J reference *GRCh38-alts-ensembl-7.1.0* and a CSV of TotalseqC antibodies for the CSP libraries were used as inputs for cellranger. Both TCR and BCR contig assemblies outputted from cellranger were further processed using Immcantation (v4.5.0), which aligned contigs to germline IMGT sequences using IgBLAST (v1.22.0). All contigs were used as input for BCR data and filtered contigs were used as input for TCR data. Clone IDs were generated using Change-O (v1.3.0) with the *aa* model. A distance threshold (*dist*) of 0.0 was used for TCR data and 0.15 for BCR data. All processed data was further analyzed using a custom bioinformatics codebase written in R (v4.5.0). Immcantation results were further filtered by only keeping contig assemblies with at least two UMIs. In addition, for each cell, the contig with the largest number of UMIs and or reads was kept. This resulted in there being one TCR alpha chain (TRA) and on TCR beta chain (TRB) per TCR as well as on immunoglobulin heavy chain (IGH) and light chain (IGK or IGL) per BCR. Gene expression (GEX) and CSP data were analyzed using Seurat (v5.5.1). On the GEX data specifically, Ambient RNA removal was performed using SoupX (v1.6.2) and doublet removal was performed using DoubletFinder (v2.0.4). Automated cell type annotation was performed on GEX data with Azimuth (v0.5.0) and used the Human PBMC atlas (*pbmcref*) as a reference. GEX and CSP Seurat objects were generated separately then overlapped. For GEX only cells with at least 400 non-zero genes were kept. For CSP, cells with at least one non-zero gene was kept. TCR V genes (TRAV, TRBV, TRGV, TRDV) were omitted from the GEX Seurat object prior to performing SCT normalization per sample. All normalized samples were then integrated on common gene anchors using Seurat’s *FindIntegrationAnchors* and *IntegrateData* commands. CAR-T cell was annotated using both the CSP and TCR V(D)J data. Specifically, both CD3 and Whitlow (CAR-T defining marker) expression were used in the CSP data. Each participant and sample were evaluated individually when defining the CAR-T cell population. Every time point outside of baseline was evaluated. For both CD3 and Whitlow, two distributions were compared. First was the distribution of raw CSP counts among cells that overlapped with TCR. The second was the raw CSP count distribution among cells that did not overlap with TCR data, which served as a background distribution. All thresholds were selected manually in relation to summary statistics such as mean and standard deviation per distribution. Each background distribution had peak towards zero, which represented signal noise. Thresholds were selected to be greater than this early peak of expression for both CD3 and Whitelow for each sample. These cells were then overlapped with the GEX data to establish the final CAR-T cell annotations. Differential gene expression (DGE) analysis was performed on the CAR-T cell population using a non-parametric Wilcoxon method built into Seurat’s *FindAllMarkers* function (method = wilcox, threshold = 0.0). Thresholds found in supplemental methods.

BCR sequencing FASTQ files were downloaded into Cellranger (10X Genomics, Pleasanton, CA) for vdj_b alignment and initial QC, then all_contig.fasta and all_contig_annotations.csv files were uploaded to PipeBio (a Benchling Company, San Francisco, CA). PipeBio analysis included identifying human heavy or light chain VDJ germline usage and identity, isotype and clonal families using IMGT v1.1. Clonal families were defined as having full identity in heavy chain germline use, CDRH1, CDRH2, CDRH3 length and entire light chain variable domain sequences with CDRH3 sequences clustered at 85% identity. Results were plotted in Excel (Microsoft, Redmond, WA).

CSF myeloid cells were selected based on Azimuth level1 annotations of monocytes. Seurat’s Aggregate Expression function was used to generate pseudobulk count matrices for each participant. EdgeR package was used for DEG analysis, normalizing library sizes using the trimmed mean of M-values. To identify differentially expressed genes in late time point (Week 24 and Week 48) compared to early (Screening and Day 14), a generalized linear model with a paired design was used. Gene set enrichment analysis was done with fsgea package using Reactome pathway gene sets

Myeloid compartment gene module scoring:

AUCell package was used to calculate per cell module scores for the following gene modules:

inflammatory_module (defined by the following genes: “S100A8”, “S100A9”, “FCN1”, “LYZ”, “CTSS”, “VCAN”, “LGALS3”, “IL1B”, “TNF”, “CCL2”, “CCL3”, “CCL4”, “TLR2”, “TLR4”, “NFKBIA”, “NFKBIZ”, “RELA”, “RELB”, “JUN”, “FOS”)

ifn_module <-c(“IFIT1”, “IFIT2”, “IFIT3”, “ISG15”, “MX1”, “MX2”, “OAS1”, “OAS2”, “OAS3”, “IFI27”, “IFI44”, “IFI44L”, “STAT1”, “STAT2”, “IRF7”, “IRF1”, “USP18)

repair_homeostatic_module <-c(“CD163”, “MRC1”, “FOLR2”, “APOE”, “TREM2”, “MSR1”, “GPNMB”, “C1QA”, “C1QB”, “C1QC”, “CX3CR1”, “SELENOP”).

### Proteomic Analysis

Sample processing and initial data analysis described in Supplemental Methods. Using an unbiased approach, we first investigated longitudinal changes in protein expression in both participants. Based on observed CAR-T and B-cell kinetics, samples were grouped into three phases: pre-CAR-T-cell therapy and lymphodepletion (“Pre”); an early post-treatment phase reflecting CAR-T-cell expansion (“Early”, Day +1 to +56); and a late post-treatment phase reflecting steady-state conditions and B-cell repopulation (“Late”, > Day +56). For each of three pairwise comparisons (Early vs. Pre, Early vs. Late, Late vs. Pre), log2 fold changes (logFC) of the differentially expressed proteins (DEPs) were estimated using *limma* (v.3.66.0) with *duplicate correlation* to account for repeated measures within participants, applying a 5% false discovery rate (FDR).

In addition, we performed Gene Set Enrichment Analysis (GSEA) using the R package *fgsea* (v.1.36.2) to assess whether specific biological pathways are up- or downregulated following CAR-T. For each comparison, all quantified proteins were ranked by ranked by their moderated t-statistic from *limma* and tested against the *Reactome* human gene sets from the *msigdbr database* (v.25.1.1) [68, 69]. Pathways were restricted to their intersection with the ∼5,400 proteins measured on the Olink panel prior to enrichment testing. Gene sets with fewer than 10 measurable proteins after this restriction were excluded. Normalized Enrichment Scores (NES) were reported together with FDR-adjusted *p*-values (5%).

Finally, we descriptively analyzed a set of predefined markers associated with B cell homeostasis (BAFF, BAFF-R, TACI, BCMA, FCRL1, FCRL2, FCER2, CD22, CXCL13, 4-1BB), microglial and myeloid activation (CHIT1, CHI3L1, CSF1R, TRME2, SPP1), neuronal or astrocyte injury (NFL, GFAP), and other immune function (IL-1A/B, IL1R1, IL-2, IL-3, IL-4, IL-5, IL-6, IL-7, IL-10, IL-12, IL-13, IL-15, IL-16, IL-17A, IL-18, IFNA1/IFNA13, IFNG, VEGF-A, CCL18, CCL2, CCL3, CD27, CXCL10, GM-CSF, TNF-alpha, granzyme A and B, and MMP9.

Methodology for Flow Cytometry, ddPCR, Recombinant Protein/Binding Assay, Luminex, PhipSeq, and Viral Association TCR Data Processing are found within Supplemental Methods.

## Data Availability

All data produced in the present study are available upon reasonable request to the authors. Data will be made available on publication.

## List of Supplementary Materials

Materials and Methods
Supplemental Figures:

S1: Clinical Outcomes
S2: Participant 1’s Adverse Event
S3: CAR-T Dynamics
S4: B cell Reconstitution
S5: B cell Homing
S6: Immunologic changes after CAR-T
S7: Vaccine Titers
Table S1: T cell response of Participant 1’s Adverse Event
References [47, 70–79]

## Acknowledgments

We would like to thank the participants and their families involved in this study as it would not have been possible without them. We would also like to thank our oncology team, dermatology, and dermatology pathology colleagues (Jeffery P. North) for their help in the care of our participants.

## Author contributions

Conceptualization: SG, MRS, RL, JW, SLH, BAC

Methodology: SG, RD, LMJ, KM, LS, JJS, AB, AA, MRW

Investigation: RD, LMJ, KM, LAS, SV, MS, RH, CC, EH

Visualization: RD, LMJ, SK, AB

Funding acquisition: SG, SLH, MRW, BAC

Project administration: RL, NO, NT, SS, SZ, CF

Writing – original draft: SG, LMJ, KM, LS, MRW, BAC

Writing – review & editing: SG, MRS, LMJ, KM, LS, RL, NO, JJJ, AA, SLH, MRW, BAC

## Data and materials availability

All data are available in the main text or the supplementary materials. Raw data will be made available upon publication: RNAseq data will be uploaded to the GEO database, PhiPSeq and proteomic data on dryad.

