## Supplemental Data for "Immune Profiling after treatment with an Anti-CD19 Chimeric Antigen Receptor T Cell Therapy in Treatment-Refractory Progressive Multiple Sclerosis"

### Supplementary Methods:

#### Study Design and Oversight

This was a phase 1, open-label, single ascending dose study of miv-cel in participants with treatment refractory progressive MS (NCT06451159). The study was designed by the lead investigators (SG, MRS, BACC). The protocol was approved by the UCSF Institutional Review Board (IRB number: 23-40657), and the study was conducted in accordance with the principles of the Declaration of Helsinki. All participants provided written informed consent. Data were collected by the investigators and interpreted by all authors. All authors participated in the preparation and approval of the final manuscript.

#### Participants:

Included participants were aged 25-70 with treatment refractory progressive MS, defined as evidence of clinical disability progression within the 2 years prior to inclusion, i.e. a) progression of EDSS during the past two years of at least 1 point sustained for at least 6 months if inclusion EDSS is from 3.5 to 5.5 or at least 0.5 point increase sustained for at least 6 months if inclusion EDSS is from 6 to 6.5 or b) increase of T25FW by at least 20% in the last two years sustained for at least 6 months or c) other well-documented objective worsening despite at least 1 year of prior treatment for progressive forms of MS (Siponimod or anti-CD20 monoclonal antibodies). Complete eligibility criteria are reported in the supplement.

#### Study Treatment

Participants underwent apheresis with target collection of $1-2 \times 10^9$ total nucleated cells (Figure 1A). Viral transduction of T-cells with the miv-cel lentivirus vector, a second generation anti-CD19 CAR with a CD28 co-stimulatory domain [70], was performed *ex vivo*. Participants were treated with lymphodepletion with intravenous fludarabine 30 mg/m<sup>2</sup> and cyclophosphamide 300 mg/m<sup>2</sup> daily from Day -5 to Day -3. Participants were then admitted to the hospital on Day -1 and received a single intravenous infusion of $0.33 \times 10^8$ CAR+ T-cells on Day 0.

#### Assessments

Participants were observed in the hospital for at least 7 days after miv-cel infusion. After discharge, participants underwent twice weekly lab and clinical monitoring up to Day 28 following miv-cel infusion. After the 28 days, participants were seen on Week 8, Week 24, Week 36, and Week 48. Neurologic exam was completed on Screening, Week 8, Week 24, Week 36, and Week 48. CSF was obtained at Screening, Day 14, Week 24 and Week 48. MRI of the brain, cervical spinal cord and thoracic spinal cord was obtained at Screening, Week 8 (MRI brain only), and Week 48. MRI sequences were acquired on a 3T Siemens Skyra Fit MRI and interpreted by a staff neuroradiologist. Patient Reported Outcomes (PROs) included Bladder/Bowel Control Scale, Modified Fatigue Scale, WHO Disability Assessment Survey, Screening assessments and EDSS were completed by a board-certified neurologist.

#### Study Objectives and Endpoints

The primary safety objective was to determine the recommended dose level of miv-cel based on incidence and severity of AE and dose-limiting toxicities. The primary efficacy objective was to assess CNS penetration of miv-cel based on presence of CAR+ T-cells in the CSF following their peak expansion in peripheral blood. Secondary objectives included characterizing target engagement within the CNS of miv-cel based on the proportion of participants in whom reduction of CNS OCB and/or normalization of CSF IgG index were detected. Exploratory objectives included efficacy of miv-cel treatment based on clinical assessments such as

EDSS, T25FW, and incidence of new MRI lesions; patient-reported outcomes; B-cell and T-cell kinetics; and other biomarkers. All endpoints were assessed by the principal investigators (SG, MS, and BCC).

##### Flow Cytometry:

Peripheral Blood Mononuclear Cells (PBMCs) were isolated from BD Vacutainer™ CPT™ Mononuclear Cell Preparation Tubes (02-688-81), treated with 1xRBC lysis buffer (Biolegend, 420301), and washed with 1% BSA in D-PBS. PBMCs were counted using the Invitrogen Countess 3 FL Automated Cell Counter and 0.04% Trypan Blue Solution (Gibco, 15250061). PBMCs were cryopreserved in 10% DMSO in FBS-HI. Cryopreserved PBMCs were flash thawed in a 32°C water bath, then washed and resuspended with 1% BSA in D-PBS. PBMCs ($1 \times 10^6$) were blocked with Human TruStain FcX™ (Fc Receptor Blocking Solution) (Biolegend 422301) and incubated at 4°C in the dark for 10 minutes. PBMCs were washed with 0.02% FBS-HI and 0.004% EDTA in D-PBS and resuspended with the following antibody panels and BD Horizon™ Brilliant Stain Buffer Plus (566385) at 4°C in the dark for 30 minutes. The B cell antibody panel included LIVE/DEAD™ Fixable Near-IR Dead Cell Stain Kit (Invitrogen L10119), CD45 Alexa Fluor 647 (Biolegend 304020), CD19 PE-cy5.5 (Biolegend 9340-16), CD3 BB515 (BD Biosciences 564465), IgD BV421 (BD Horizon 562518), CD24 PE/Dazzle 594 (Biolegend 311134), CD27 PE (Biolegend 302807), and CD38 PE-cy7 (Biolegend 303516). After staining, PBMCs were washed and resuspended in 0.02% FBS-HI and 0.004% EDTA in D-PBS. PBMCs were then analyzed on the BD LSRFortessa™.

##### Digital Droplet PCR:

DNA was extracted from participant's isolated PBMCs (Zymo Research Quick-DNA/RNA MicroPrep Plus) according to manufacturer's instructions. DNA was eluted in 30uL of nuclease-free water, and the DNA concentration was measured using a NanoDrop ND-1000 spectrophotometer.

Based on a previously published protocol [71], the digest mix contained the following: 3.65uL nuclease-free water, 1uL 10x NEB Buffer, 0.1uL 2% BSA, and 0.25uL DraI restriction enzyme. Digest mix was added at a 1:1 ratio with DNA (10uL sample, 10uL digest). Digest was done simultaneously with PCR using the cycling conditions.

Primers and probe sequences were provided from Kyverna and ordered from IDT with primers at a 25nM concentration and probe at 250nM concentration with a 5' FAM for KYV101 and 5' HEX for RPP30. Forward primer KYV101: ACCGGTCCAGAGTGAAGTTCA, Reverse Primer KYV101: GCTCGTTGTACAGCTGGTTCTGT, Probe KYV101: CAGATCCGCCGACGC, Forward primer RPP30: GAAGAAACCTCGGCCATCAG, Reverse primer RPP30: CCCTCACACTTGGCTTTCTT, Probe RPP30: AGGAGATGAAGATTGTCTTCCAGCTTCC.

DdPCR master mix was made in duplicate containing 12.5uL ddPCR supermix for probes, 1.25uL 20x primer-probe mix for KYV101, 1.25uL 20x primer-probe mix for RPP30 (100uL of 20x primer-probe mix created by adding 18uL 100uM forward primer, 18uL 100uM reverse primer, 5uL 100uM probe, and nuclease-free water added to bring total volume up to 100uL). Following this, 30uL of master mix was aliquoted into a PCR tube and 20uL DNA + digest mix was added. All reagents were vortexed well before droplet generation.

In a DG8 cartridge, 20uL of the DNA ddPCR mixture was added into each sample well, and 70uL of droplet generation oil for probes was added into the oil well. Once droplets were generated, they were dispensed into ddPCR 96 well Semi-Skirted plates. The plate was heat sealed with foil and placed into the thermocycler. The thermocycler was set with a 2°C/s ramp rate with the following cycling conditions: 98°C 10min for 1 cycle, 94°C 30sec for 40 cycles, 60°C 60sec for 40 cycles, 98°C 10 min for 1 cycle, 4°C hold. Plate was then placed into a QX 200 Droplet Reader for probe detection.

##### Single cell RNA sequencing with Feature Barcode Technology:

Fresh CSF was centrifuged at 4°C in 15mL conical tubes (Falcon) and supernatant was removed. The remaining cell pellet was resuspended with 1% BSA in D-PBS. PBMCs were isolated from whole blood collected in CPT tubes (BD), treated with 1x RBC lysis buffer (Biolegend, 420301), and washed twice with 1% BSA in D-PBS. CSF cell and PBMC suspensions were blocked with Human TruStain FcX™ (Fc Receptor Blocking Solution) (Biolegend 422301) and incubated at 4°C for 10 minutes. Cell suspensions were then incubated with TotalSeq™-C Human TBNK Cocktail (Biolegend 399903) per manufacturing instructions at 4°C for 30 minutes. TotalSeq-C0100 anti-human CD20 Antibody (Biolegend 302363), TotalSeq™-C 0154 anti-human CD27 (Biolegend 302853), TotalSeq™-C0410 anti-human CD38 Antibody (Clone: HB-7) (Biolegend 356637), and Whitlow/218 Linker (E3U7Q) Rabbit Monoclonal Antibody (Cell Signaling Technology #66159) TotalSeq™-C 5078 Custom Oligo Conjugation (Biolegend) were included at concentrations scaled by optimal titration results. Cell suspensions were washed with 1% BSA in D-PBS, 0.04% BSA in D-PBS, and resuspended. Cells were counted with 0.04% Trypan Blue Solution (Gibco, 15250061) on a hemocytometer. CSF cell and PBMC suspensions were immediately run on the 10x Genomics Chromium iX to form droplets and generate barcoded cDNA. Gene expression, Cell Surface Protein, and BCR and TCR enriched V(D)J libraries were prepared according to the Chromium GEM-X Single Cell 5' Reagent Kits v3 with Feature Barcode technology for Cell Surface Protein (10x Genomics, CG000734) kit user guide. Library quality was assessed using High Sensitivity D5000 ScreenTape Analysis (Agilent) and quantified using Lib Quant Kit (Illumina/Uni) (Roche, 07960140001). Sequencing was performed on the NovaSeq X (Illumina) with NovaSeq™ X Series 25B Reagent Kit (100 Cyc) 100-cycle Kit (Illumina, 20125967). Libraries were sequenced at a minimum target depth of 20,000 read pairs/cell for 5' Gene Expression libraries and 5,000 read pairs/cell for V(D)J and Cell Surface Protein libraries.

##### CITE-Seq Custom Barcode Antibody

Whitlow/218 Linker (E3U7Q) Rabbit Monoclonal Antibody (Cell Signaling Technology #66159) was sent to Biolegend for TotalSeq™-C 5078 Custom Oligo Conjugation. To determine the optimal concentration of Feature barcoded antibodies to use with Chromium GEM-X Single Cell 5' Reagent Kits v3 with Cell Surface Protein, a six-point titration was performed. PBMCs were incubated with PE anti-human CD20 Antibody (Biolegend 302305), PE anti-human CD27 Antibody (Biolegend 356405), PE/Cyanine7 anti-human CD38 Antibody (Biolegend 356607), and Whitlow/218 Linker (E3U7Q) Rabbit Monoclonal Antibody (PE Conjugate) (Cell Signaling Technology #62405) and analyzed on the BD LSRFortessa™. The optimal volume of antibody stock needed for TotalSeq™-C antibody spike-ins was then be calculated for the cell staining.

##### Single Cell Sequencing Sample Processing:

Fresh CSF was centrifuged at 4°C in 15mL conical tubes (Falcon), and supernatant was removed. The remaining cell pellet was resuspended with 1% BSA in D-PBS. PBMCs were isolated from whole blood collected in CPT tubes (BD), treated with 1x RBC lysis buffer (Biolegend, 420301), and washed twice with 1% BSA in D-PBS. CSF cell and PBMC suspensions were blocked with Human TruStain FcX™ (Fc Receptor

Blocking Solution) (Biolegend 422301) and incubated at 4°C for 10 minutes. Cell suspensions were then incubated with TotalSeq™-C Human TBNK Cocktail (Biolegend 399903) per manufacturing instructions at 4°C for 30 minutes. TotalSeq-C0100 anti-human CD20 Antibody (Biolegend 302363), TotalSeq™-C 0154 anti-human CD27 (Biolegend 302853), TotalSeq™-C0410 anti-human CD38 Antibody (Clone: HB-7) (Biolegend 356637), and Whitlow/218 Linker (E3U7Q) Rabbit Monoclonal Antibody (Cell Signaling Technology #66159) TotalSeq™-C 5078 Custom Oligo Conjugation (Biolegend) were included in the incubation at concentrations scaled based on optimal titration results. Antibody titrations were performed using stained PBMCs with PE anti-human CD20 Antibody (Biolegend 302305), PE anti-human CD27 Antibody (Biolegend 356405), PE/Cyanine7 anti-human CD38 Antibody (Biolegend 356607), and Whitlow/218 Linker (E3U7Q) Rabbit Monoclonal Antibody (PE Conjugate) (Cell Signaling Technology #62405) on the BD LSRFortessa™. Cell suspensions were washed with 1% BSA in D-PBS, 0.04% BSA in D-PBS, and resuspended.

Cells were counted with 0.04% Trypan Blue Solution (Gibco, 15250061) on a hemocytometer. CSF cell and PBMC suspensions were immediately run on the 10x Genomics Chromium iX to form droplets and generate barcoded cDNA. Gene expression, Cell Surface Protein, and BCR and TCR enriched V(D)J libraries were prepared according to the Chromium GEM-X Single Cell 5' Reagent Kits v3 with Feature Barcode technology for Cell Surface Protein (10x Genomics, CG000734) kit user guide. Library quality was assessed using High Sensitivity D5000 ScreenTape Analysis (Agilent) and quantified using Lib Quant Kit (Illumina/Uni) (Roche, 07960140001). Sequencing was performed on the NovaSeq X (Illumina) with NovaSeq™ X Series 25B Reagent Kit (100 Cycles) 100-cycle Kit (Illumina, 20125967). Libraries were sequenced at a minimum target depth of 20,000 read pairs/cell for 5' Gene Expression libraries and 5,000 read pairs/cell for V(D)J and Cell Surface Protein libraries.

##### Threshold for defining CAR-Ts:

| Participant | timepoint | CD3_cutoff | Whitlow_cutoff |
| --- | --- | --- | --- |
| **CART0001** | D14 | 80 | 15 |
| **CART0001** | AE2 | 300 | 200 |
| **CART0001** | W8 | 100 | 20 |
| **CART0001** | W24 | 200 | 80 |
| **CART0001** | W48 | 300 | 70 |
| **CART0002** | D14 | 200 | 100 |
| **CART0002** | W8 | 100 | 20 |
| **CART0002** | W24 | 150 | 50 |
| **CART0002** | W48 | 150 | 50 |

##### Proteomic Data Sample and Data Processing:

Serum samples were collected longitudinally and stored at -80 °C until processing. Proximity extension assays (PEA) were performed using the Olink Explore HT panel (Olink®) as described previously [72]. Briefly, this approach combines antibody-based protein detection with next-generation sequencing (NGS) to measure >

5,400 proteins. Relative protein expression was quantified as normalized protein expression (NPX) values, an arbitrary log<sub>2</sub>-scale in relation to plate controls. To adjust for inter-plate variability across multiple runs, we used plate control-normalized NPX values, which are calibrated using inter-plate control samples included on every plate. Quality control (QC) was conducted at both the sample and assay (= protein) levels [72].

Data processing and statistical analyses were performed using R v.4.5.2 (2025-10-31), R Studio v.2026.1.1.403, and the R packages *OlinkAnalyze* (v.4.5.0, Nevola K. *OlinkAnalyze*: facilitate analysis of proteomic data from Olink. CRAN. Accessed December 1, 2025. https://CRAN.R-project.org/package=OlinkAnalyze), *limma* (v.3.66.0) [73], and *fgsea* (v.1.36.2). Sample outliers were identified using principal component analysis via *olink\_pca\_plot()*, defined as samples exceeding $\pm 3$ standard deviations on either PC1 or PC2. Of 32 collected serum samples, only one sample showed sample-level QC warnings and was excluded from the analysis. Day 0 samples (n = 1 per participant) and samples obtained during the AE in participant 1 (n = 3) were retained for longitudinal visualization only. Among 27 serum samples included in the statistical analysis (n = 13 from participant 1; n = 14 from participant 2), only 7 of 5,401 assays were excluded because of assay-level QC warnings. Of 10 collected CSF samples, 9 were included in downstream analyses (n = 4 from participant 1, n = 5 from participant 2); one sample obtained during the AE in participant 1 was used for descriptive analysis only. In the CSF dataset, one assay block (~580 assays) from a single sample (participant 1, Week 48) failed QC and was excluded, along with two additional assays flagged with QC warnings.

#### Recombinant Proteins and Binding Assays

In house recombinant GST-EBNA1 (UniProt P03211, amino acids 386-500) was produced as previously described (M<sup>c</sup>Cutcheon *et al*, 2026) and full-length recombinant HIS-EBNA1 purchased from Abcam (UniProt P03211, Waltham, MA). After coating 100 μL/ well of 2 μg/mL recombinant EBNA1 antigens in PBS, pH7.2 overnight at 4°C in 96-well Nunc Maxisorp<sup>®</sup> plates (Electron Microscopy Sciences, Hatfield, PA), plates were washed 3x PBST and blocked for 1h in 1%BSA/PBS. Block buffer was removed and serum titrated 1/3 in duplicate, starting from 1/50 in 0.5%BSA/PBST. Serum was allowed to bind for 2h at ambient temperature, plates were washed 3x PBST and 1/5000 each of anti-kappa and anti-lambda light chain HRP antibodies (Southern Biotech, Birmingham, AL) was added in 0.5%BSA/PBST for 1h at ambient temperature. After a final 5x PBST wash plates were developed with SuperBlue TMB (Seracare Life Sciences Inc, Milford, MA). Titration results were plotted in GraphPad Prism (Boston, MA).

#### Luminex Assay

IgG directed to the EBV epitope EBNA1<sub>393-412</sub> was measured with BSA-conjugated peptide (Lifetein LLC, Somerset, NJ) or BSA alone coupled to magnetic Luminex xMAP<sup>®</sup> beads (Luminex Corp., Austin TX). Serum diluted 1/250 or CSF diluted 1/40 was quantitatively measured as BSA normalized mean fluorescence intensity (MFI) compared to sera from healthy controls (n=91 HC) as described [74]. A monoclonal antibody to the peptide [75], generated recombinantly in house, was used as a control in the assay at 0.5 ng/mL.

#### Phage Immunoprecipitation Sequencing (PhIP-Seq):

PhIP-Seq was performed using a multichannel protocol adapted from our previously published workflow [47]: (https://www.protocols.io/view/derisi-lab-phage-immunoprecipitation-sequencing-ph-czw7x7hn?step=14.1)

The human peptidome library, described previously [76], is a custom T7 bacteriophage display collection of ~731,724 unique clones, each displaying a distinct 49–amino acid peptide on the phage surface. Collectively these peptides tile the annotated human proteome with 25–amino acid overlaps between adjacent fragments. For each assay, 1 mL of phage library was incubated overnight at 4 °C with 1 µL of human serum or 20 µL of human CSF. Antibody-bound phage were then immunoprecipitated using 25 µL of a 1:1 mixture of protein A and protein G conjugated magnetic beads (Thermo Fisher Scientific, Waltham, MA; #10008D and #10009D). Following washing, bead-bound phage were eluted into 1 mL of *E. coli* BLT5403 (EMD Millipore, Burlington, MA) at an OD600 of 0.5–0.7 and amplified at 37 °C. The amplified phage pool was subjected to a second round of enrichment using samples from the same individual, with the immunoprecipitation and amplification steps repeated as above. Phage DNA was then isolated from the twice-enriched pool, barcoded and PCR-amplified, and prepared with Illumina sequencing adapters. Libraries were sequenced on an Illumina platform (Illumina, San Diego, CA) to a target depth of approximately 1 million reads per sample.

As described previously [77], amino acid–level alignment of next-generation sequencing reads from fastq files was carried out with RAPSearch2 [78]. Analysis of the human peptidome was conducted at the protein level throughout, with all reads from a given protein-coding-sequence summed. To account for differences in sequencing depth, reads from each sample were normalized as percent of total reads.

#### TCR Sequencing Data Processing and Identification of Virus-Associated TCRs

Single-cell T cell receptor (TCR) sequencing data were obtained from peripheral blood mononuclear cells (PBMCs) and cerebrospinal fluid (CSF) collected from two participant with multiple sclerosis (MS) who underwent CD19 CAR-T cell therapy. Samples were collected at five timepoints: before treatment (screening), and at day 14 (D14), week 8 (W8), week 24 (W24), and week 48 (W48) after treatment. Pre-processed single-cell TCR sequencing data were provided and used for downstream analysis. Virus-associated TCRs were identified by exact CDR3 amino acid sequence matching of TRB chains against the VDJdb database (accessed December 2025). Only human TRB entries with a VDJdb confidence score $\geq 2$ (high confidence: specificity verified with good TCR sequence confidence) were included. The following viruses were queried: Epstein-Barr virus (EBV), cytomegalovirus (CMV), human herpesvirus (HHV), herpes simplex virus (HSV-1/2), varicella-zoster virus (VZV), influenza A/B, and SARS-CoV-2. Virus-associated TCR frequency was calculated as the proportion of virus-specific cells among total T cells within each participant-compartment-timepoint combination. Virus-associated TCR frequencies were compared before and after CAR-T therapy. The post-treatment frequency was defined as the mean across D14, W8, W24, and W48. Due to the small sample size ($n=2$), formal statistical testing was not performed, and results are presented descriptively with individual participant data and mean trends. All analyses were performed in R using the *alakazam*, *dplyr*, *tidyr*, and *ggplot2* packages.

### SUPPLEMENTAL FIGURES:

**Supplemental Figure 1: Clinical Outcomes** **A.** Component of composite score. (left) Speed walking test for a distance of 25 feet, averaged over 2 trials without assistance. (middle) PASAT testing, with random versions given to prevent learning effects. (right) 9-Hole Peg Test: Speed testing of both hands to place and remove 9 pegs. **D.** Compositae of longitudinal patient reported outcomes: WHODAS, bowel/bladder, and fatigue.

**Supplemental Figure 2: Participant 1's Adverse Event.** **A.** Dermatologic punch biopsy of L chest epidermis, post H&E and staining for B and T cells showing disruption of epidermal layers by sparse T cells. **B.** Participant 1's Day 48 CAR-T proportion in PBMC and CSF via single cell analysis and T cell subtypes based on single cell gene expression of CAR+ (top) and non-CAR (bottom) cells **C.** Single cell TCR repertoire analysis comparing D14 (peak CAR-T) to adverse event (Day 48) looking for compartmental shifts and shared clonotypes over timepoints.

**Supplemental Figure 3: CAR-T Dynamics:** **A.** Participant 1's and **B.** Participant's 2 Day 14 T cell subtypes based on single cell gene expression of non-CAR cells. **C.** Participant 1's and **D.** Participant's 2 single cell TCR repertoire analysis comparing D14 (peak CAR-T) to screening looking for compartmental shifts and shared clonotypes over timepoints. **E.** Gene expression level and significance of a pre-selected list of single cell gene expression list exploring T cell subtypes and states from CAR+ T cells at Day 14 (peak CAR-T expansion). **F.** Participant 1's and **G.** Participant's 2 Day 14 CAR-T cell vs Screening T cell trafficking markers (CSF assay failure from Screening for Participant 2) based on PBMC or CSF. **H.** Reactome Pathways Analysis of Differential Gene Expression from Figure 2G, CAR+ in CSF vs peripheral blood. **I.** Differential Gene Expression between CAR+ vs non-CAR cells in the CSF. Red dots are significant with a p value < 0.05.

**Supplemental Figure 4: B cell Reconstitution.** **A.** Flow cytometry analysis from week 24 peripheral blood looking at lymphocytes => single cell => live cells. Defined B cells as CD45+CD3-CD19+. From there looked at CD24hiCD38hi and IgD+CD27-. **B.** Single Cell BCR repertoires and clonal abundance in PBMCs vs CSF and from week 24 to 48 per participant. **C.** The Immunoglobulin heavy chain (left) or light chain (right) CDR3 length distributions in participant 1 or 2 at weeks 24 or 48 from 10X Genomics sample processing. Data for participant 1 CSF at week 48 is also shown. Other times points had insufficient B cell counts to analyze. The darker the shading the greater the percentage of BCR with that CDR3 length. **D.** The isotypes of the immunoglobulin heavy (in gene locus class switching order: IgM, IgD, IgG3, IgG1, IgA1 and IgA2) and light chain (kappa or lambda) BCRs in participant 1 or 2 at weeks 24 or 48 from 10X Genomics sample processing. Data for participant 1 CSF at week 48 is also shown. Other times points had insufficient B cell counts to analyze. The cell numbers are shown in the boxes with darker shading representing greater proportions of BCR. **E.** The Immunoglobulin heavy chain variable gene (left) or light chain variable gene (right) usage in participant 1 or 2 at weeks 24 or 48 from 10X Genomics sample processing. Data for participant 1 CSF at week 48 is also shown. Bubble size is % of all 9,678 sequences. Other times points had insufficient B cell counts to analyze. The larger the bubble the greater the percentage of BCR from that gene family.

**Supplemental Figure 5: B cell homing.** **A.** Longitudinal normalized protein expression (NPX) levels of B cell markers, measured by proximity extension assays, in serum (blue) and CSF (red) from participant 1 (filled circle) and participant 2 (open square). For clarity, the early post-CAR-T period (day +1 to +56) is expanded approximately threefold on the x-axis relative to the pre-CAR-T and late post-CAR-T periods. The vertical dashed line at day +84 indicates the time of B-cell reconstitution. For FCRL1, one CSF data point from participant 1 was excluded following assay-specific quality control. BAFF, B-cell activating factor; BCMA, B-cell maturation antigen; FCER2, Fc epsilon receptor II; FCRL1/2, Fc receptor-like 1/2. **B.** Gene expression level and significance of a pre-selected list of single cell gene expression list exploring B cell maturation and survival genes from Week 24 and Week 48, combined analysis for both participants.

**Supplemental Figure 6:** Immunologic changes after CAR-T. **A.** Longitudinal single cells based UMPAS combining participants from Screening to Week 48. **B.** Single cell TCR repertoire analysis comparing Screening to Week 48 looking for compartmental shifts and shared clonotypes over timepoints. **C.** Participant 1's and **D.** Participant's 2 gene expression level and significance of a pre-selected list of single cell gene expression list exploring T cell subtypes. **E.** Gene-expression levels of T follicular helper cell markers in PBMC and CSF for each participant. **F.** Frequency of virus-associated T cells (% of total T cells) in PBMC and CSF before (screening) and after (day 14, Week 8, Week 24, and Week 48) CAR-T therapy. Virus-associated T cells were identified by exact CDR3 amino acid sequence matching of TRB chains against the VDJdb database (score $\geq 2$). The dashed line indicates the mean of both participants. EBV, Epstein-Barr virus; CMV, cytomegalovirus; SARS-CoV-2, severe acute respiratory syndrome coronavirus 2. **H.** Proteomic changes in serum and CSF across the pre-CAR-T, early post-CAR-T, and late post-CAR-T periods. Volcano plots showing differentially expressed proteins (DEPs) between the early post-CAR-T (day +1 to +56, $n = 15$ samples) and pre-CAR-T period ($n = 6$ samples, left), the late post-CAR-T ($> \text{day } +56$, $n = 6$ samples) and pre-CAR-T period (middle), and the late versus early post-CAR-T period (right). Comparisons were performed using limma. Significant proteins at FDR of 5% and 10% are highlighted in red and orange, respectively. The top 35 DEPs, ranked by $\text{padj}$, are labeled. Vertical dashed lines indicate a $\log_2$ fold-change threshold of 1, and the horizontal dashed line denotes the nominal significance cutoff ($p_{\text{nom}} = 0.05$). **I.** Longitudinal normalized protein expression (NPX) levels of markers associated with microglial and myeloid activation, neuronal and astrocytic injury, and immune function, measured by proximity extension assays, in serum (blue) and CSF (red) from participant 1 (filled circle) and participant 2 (open square). For clarity, the early post-CAR-T period (day +1 to +56) is expanded approximately threefold on the x-axis relative to the pre-CAR-T and later post-CAR-T intervals. For TREM2, CD27, CCL2, CXCL10, and IL-18, one CSF data point from participant 1 was excluded following assay-specific quality control.

**Supplemental Figure 7.** CLIA certified vaccine titers from screening to week 48 (EOS). Participants were not revaccinated against the titers measured.

SUPPLEMENTAL FIGURES:

Supplemental Figure 1:

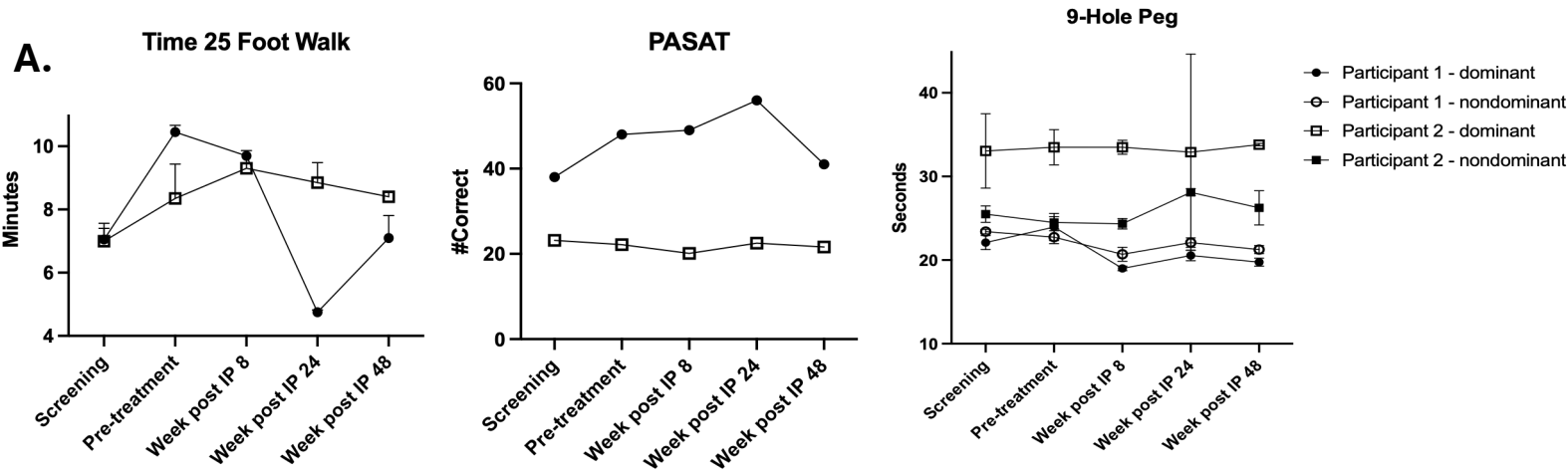

**B. Patient Reported Outcomes**

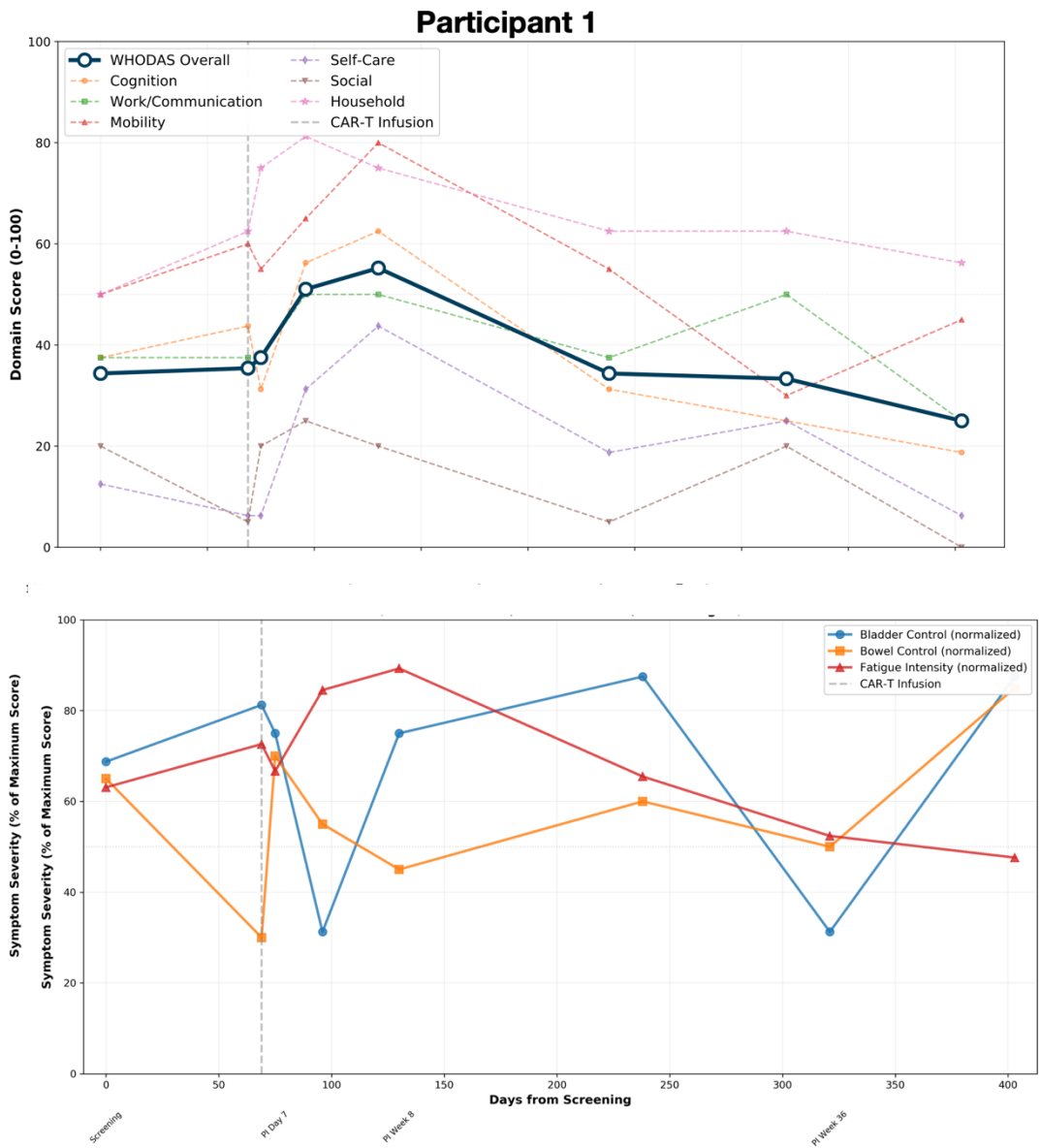

### Participant 2

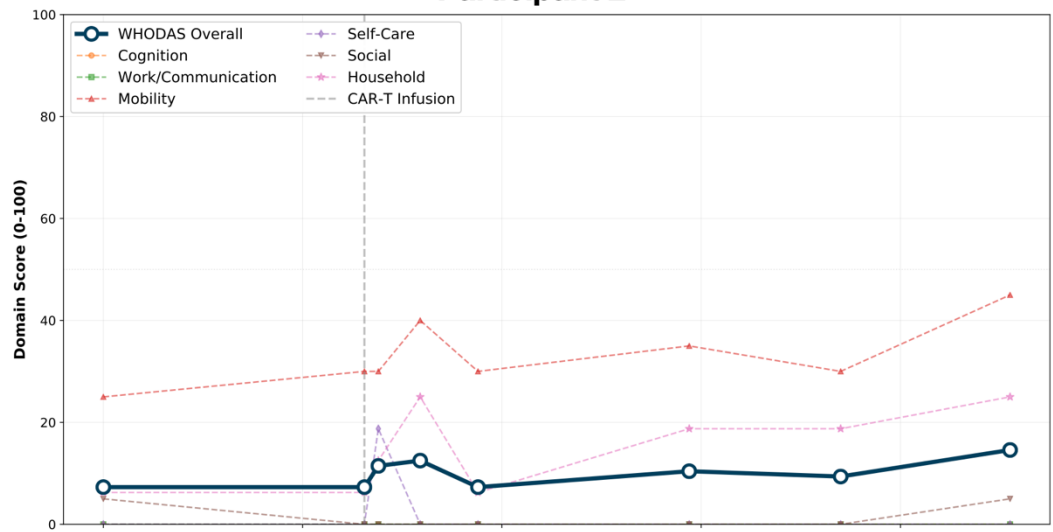

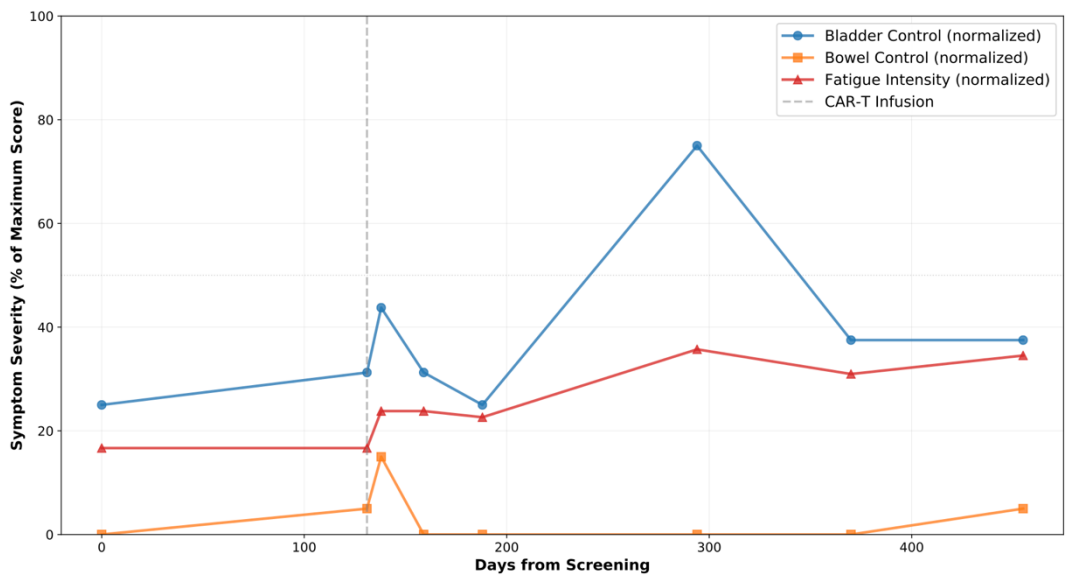

Supplemental Figure 2: Participant 1's Adverse Event

A.

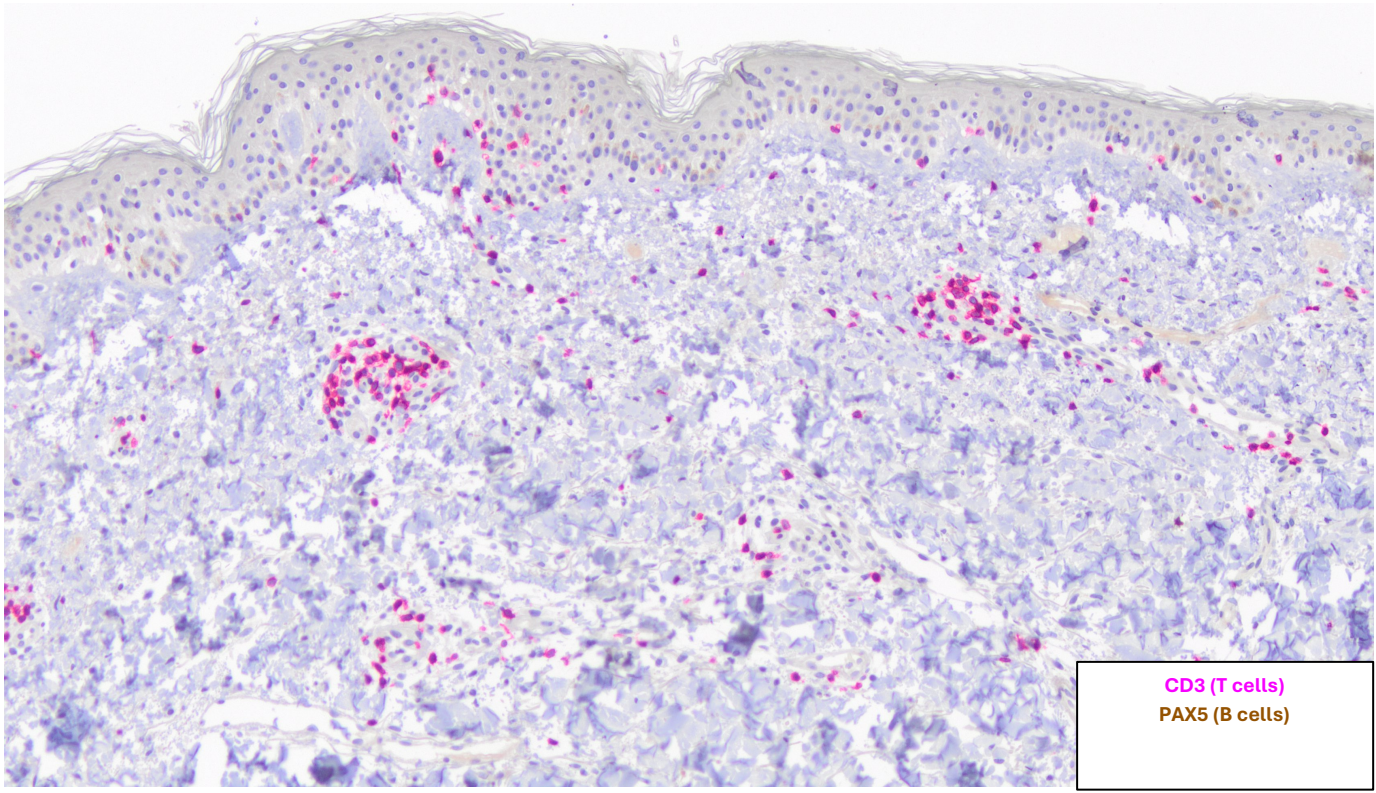

B.

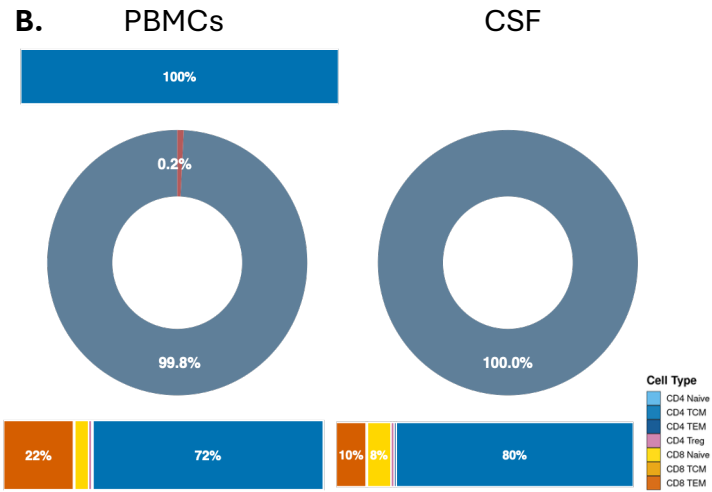

C.

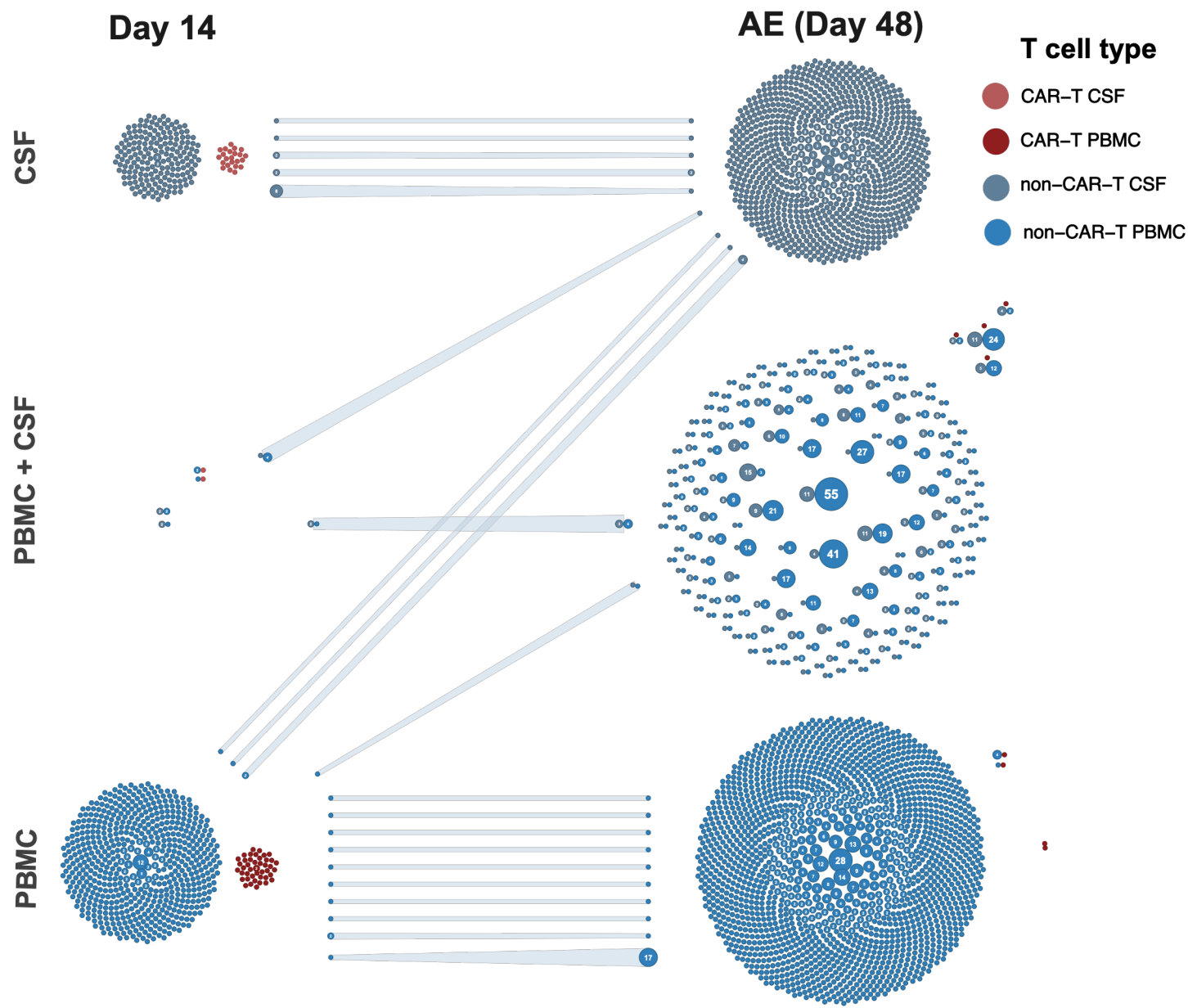

**Supplemental Figure 3: Non-CAR-T cells at Day 14**

**A. Participant 1: Non-CAR-T cells at Day 14**

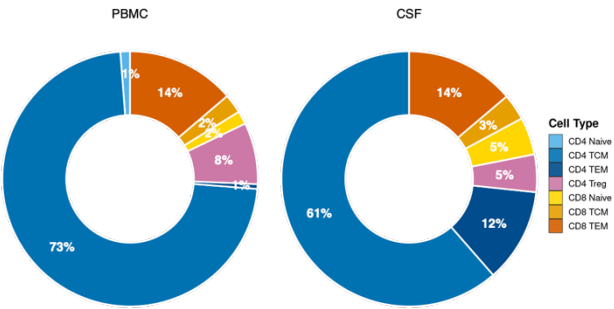

**B. Participant 2: Non-CAR-T cells at Day 14**

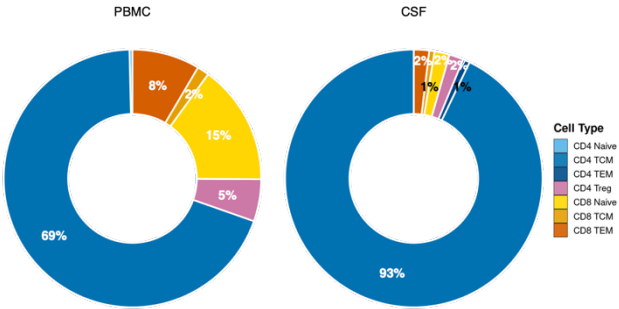

**C. Participant 1: TCR repertoire from screening to D14**

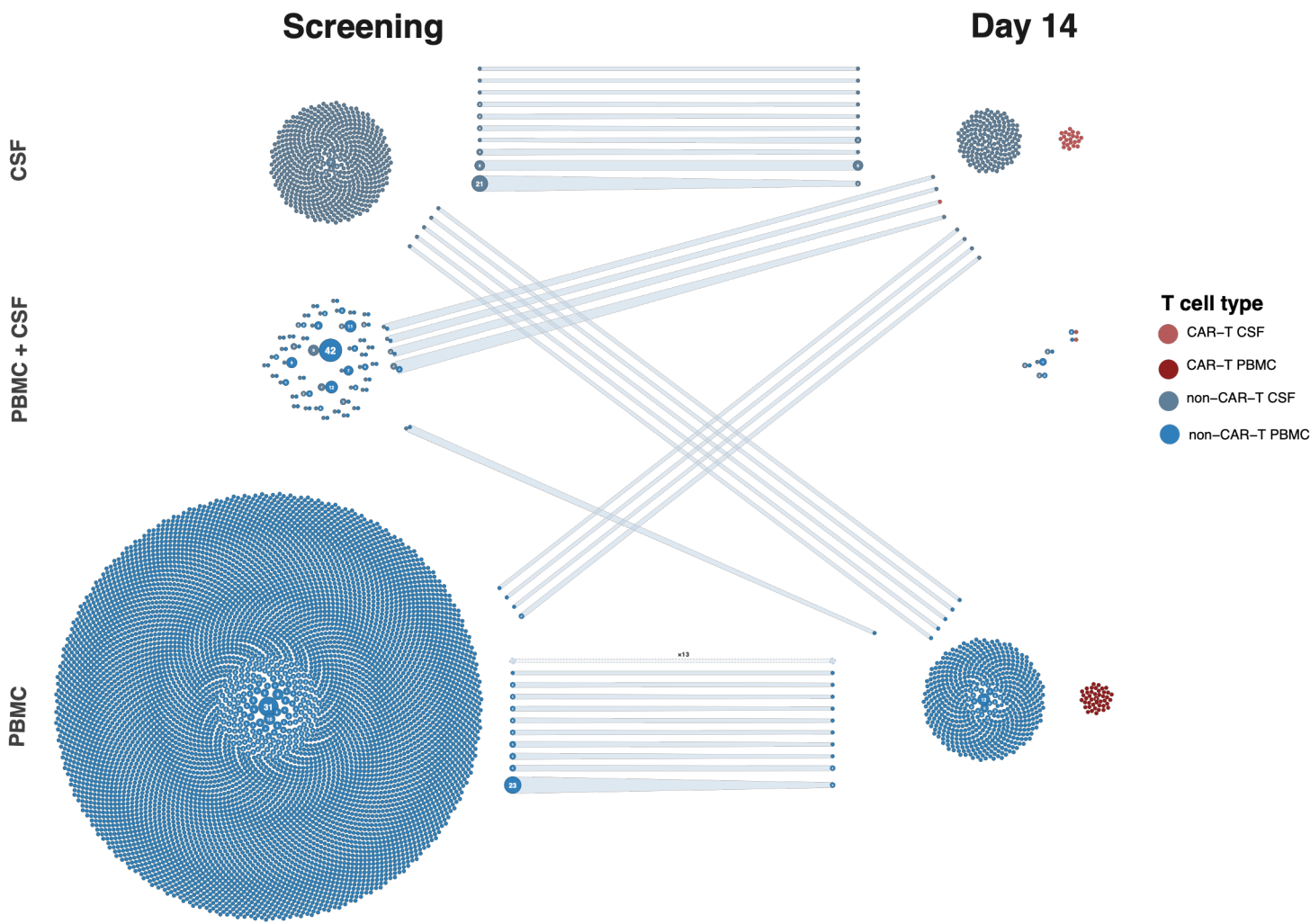

D. Participant 2: TCR repertoire from screening to D14 (CSF assay failure at screening)

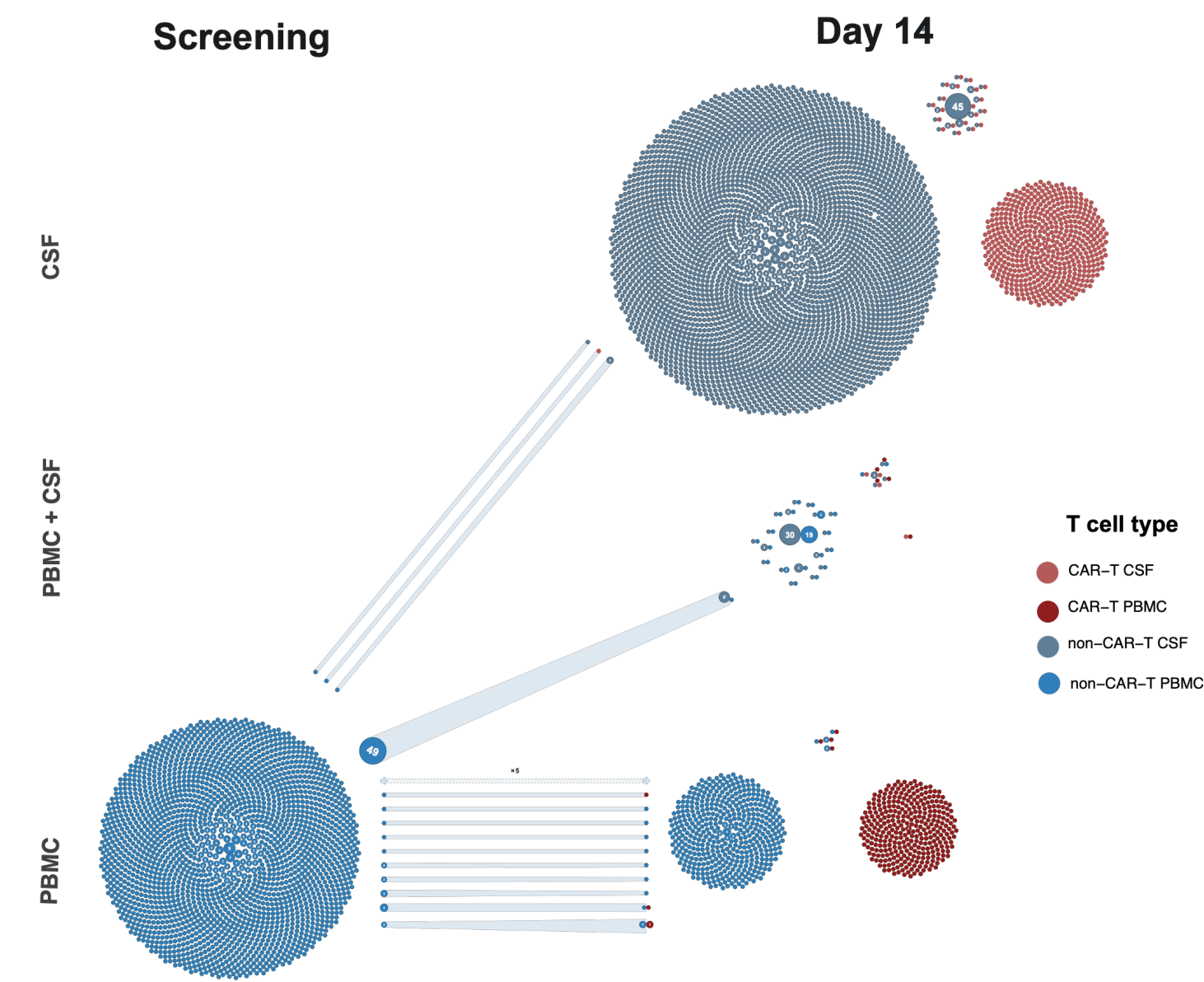

E.

Participant 1

Participant 2

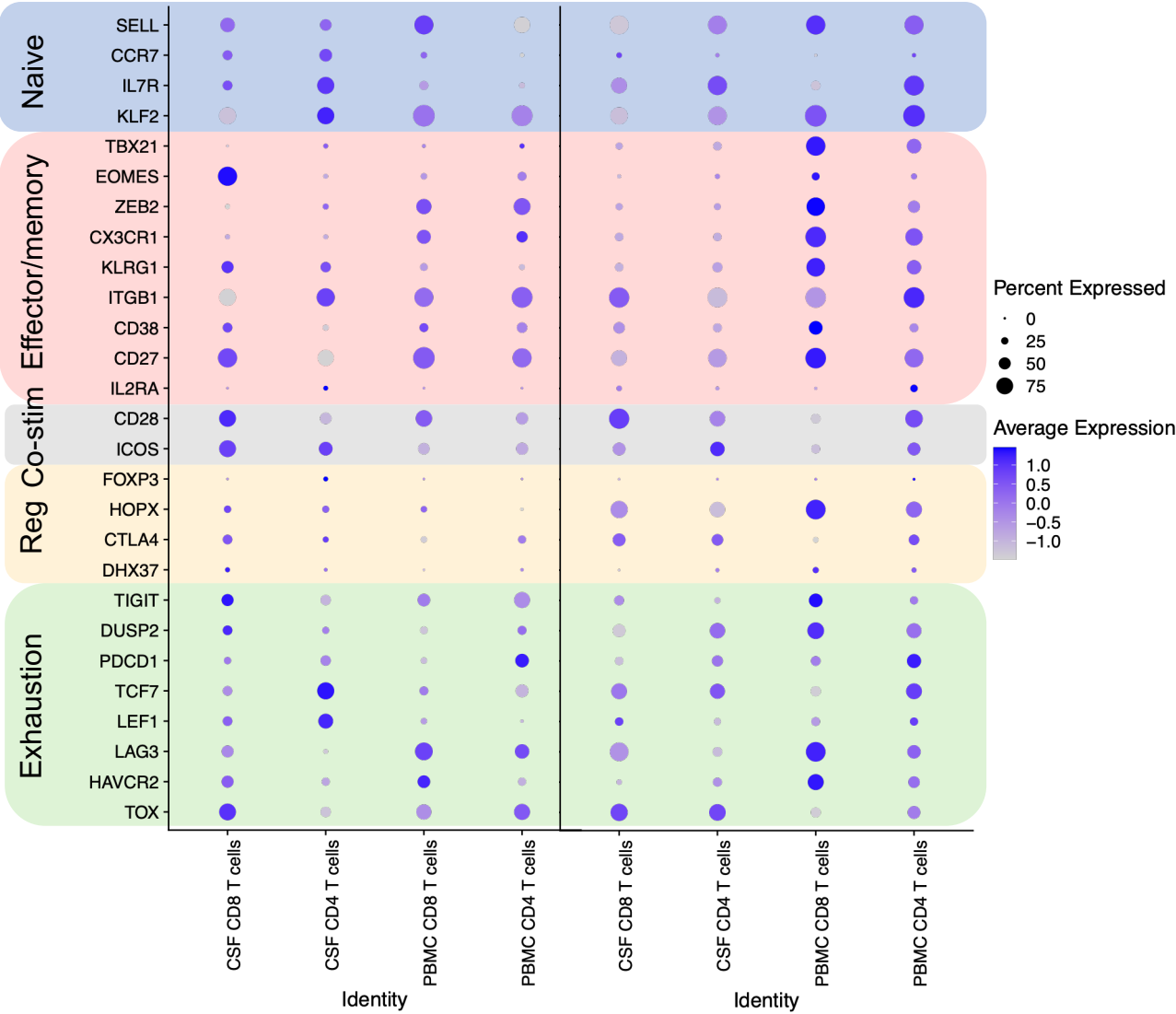

F. Participant 1: CAR-T cells at Day 14 vs Screening all T cells, Trafficking Markers

Day 14 Screening

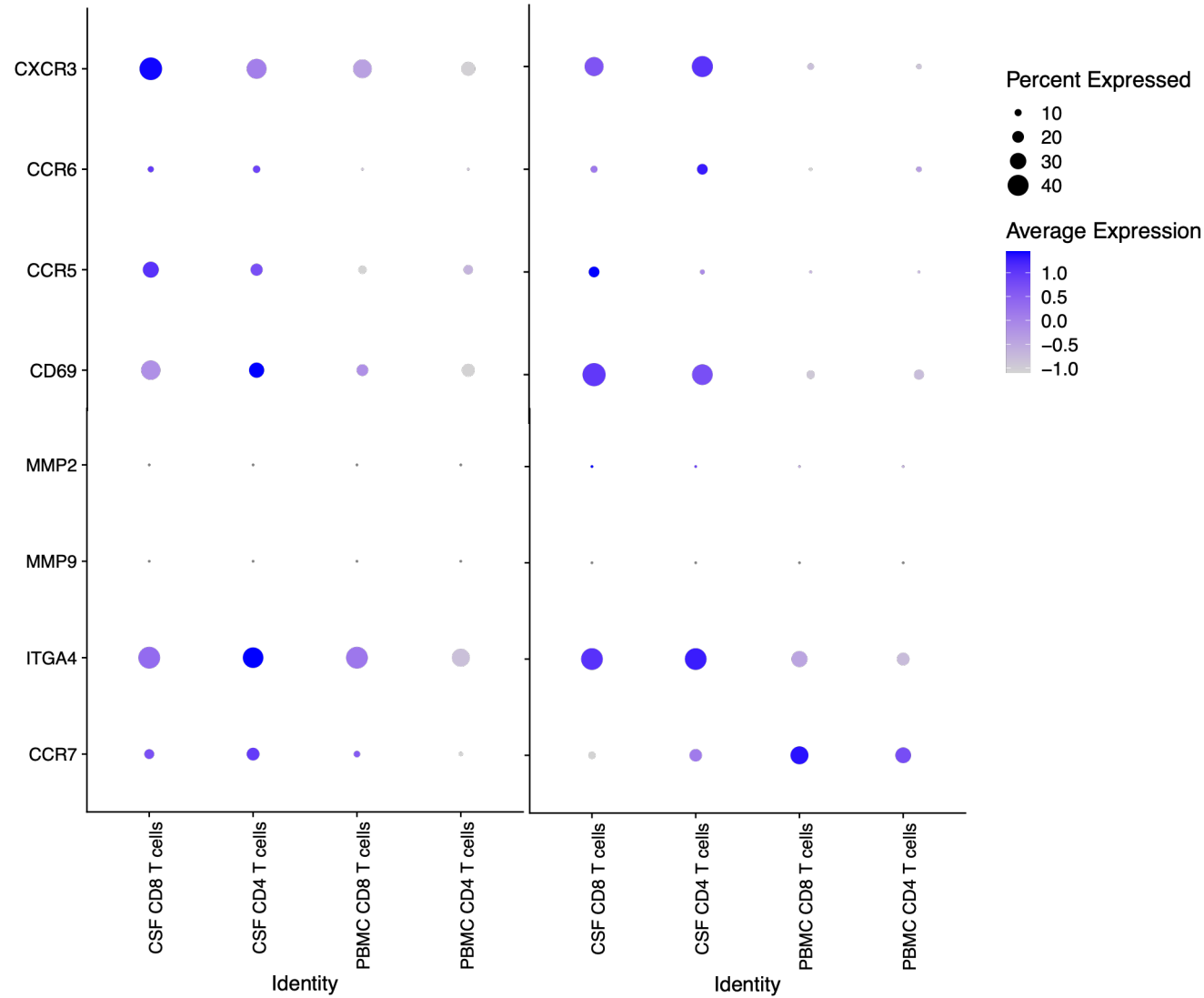

G. Participant 2: CAR-T cells at Day 14 vs Screening all T cells, Trafficking Markers (CSF assay failure at Screening)

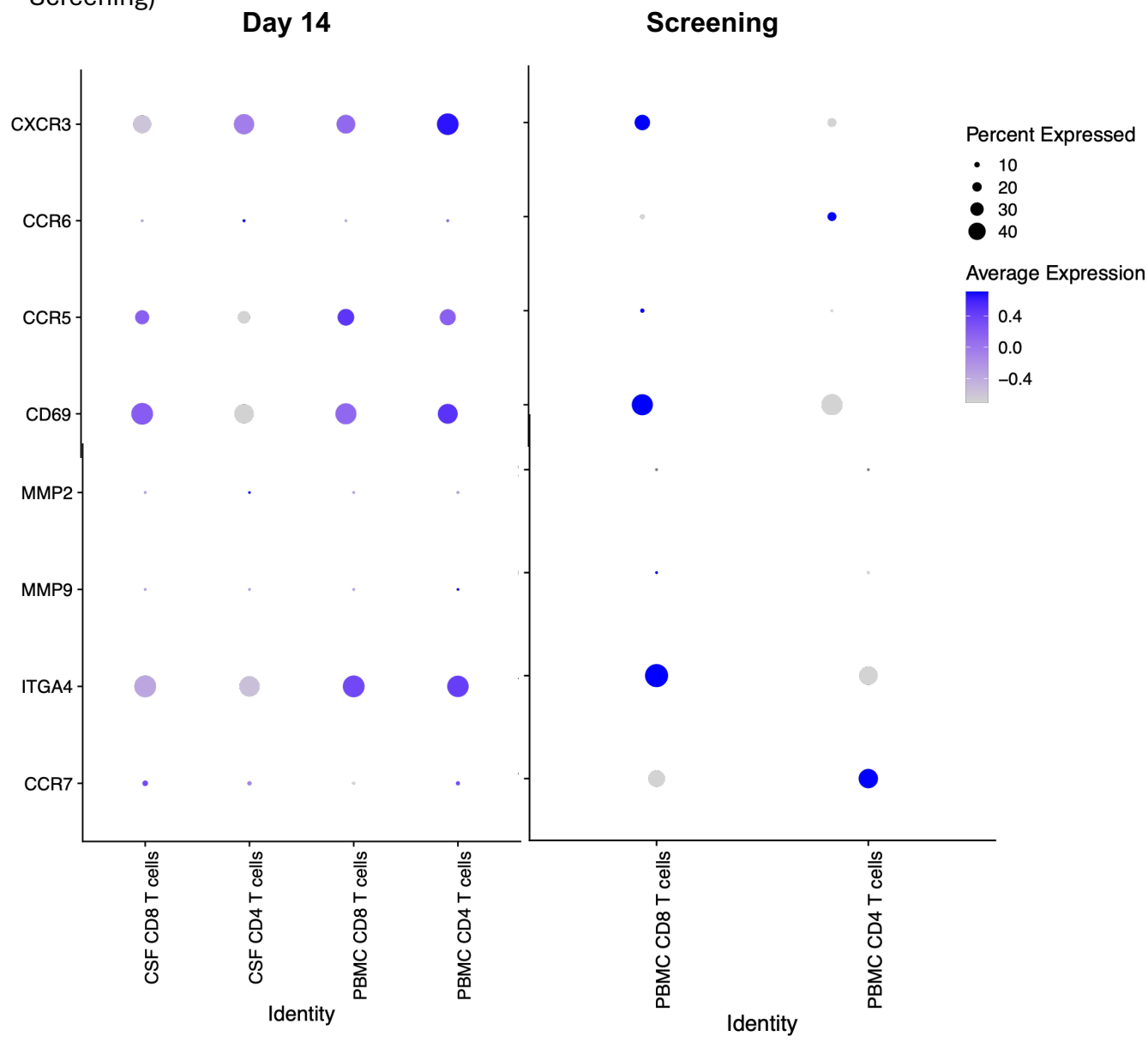

H. Reactome Pathways Analysis of Differential Gene Expression from Figure 2G, CAR+ in CSF vs peripheral blood

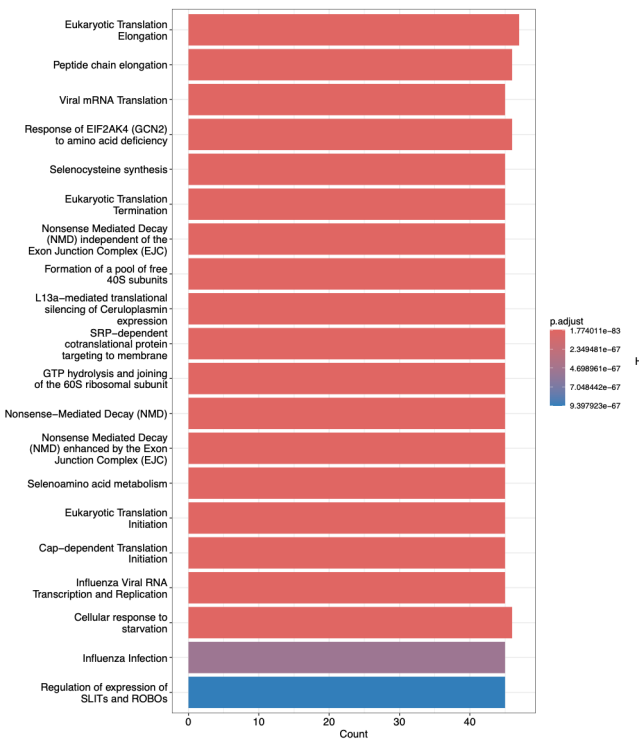

I. Differential Gene Expression between CAR+ vs non-CAR cells in the CSF

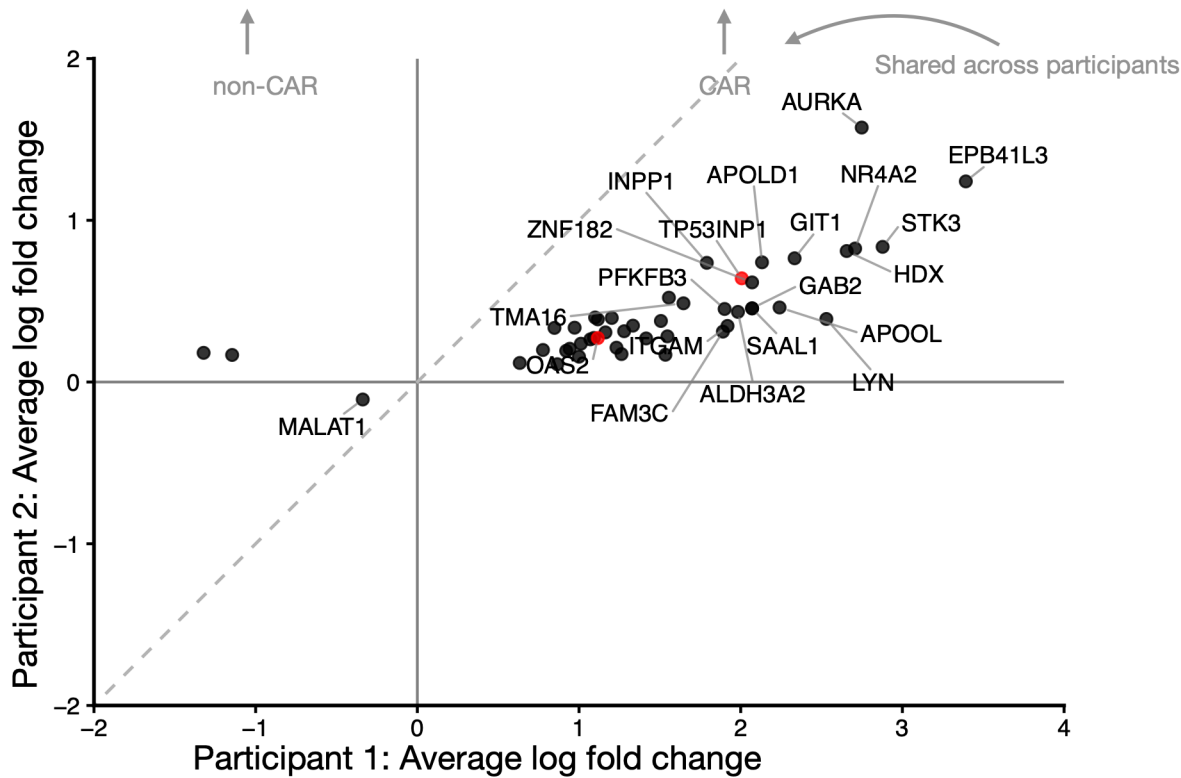

Supplemental Figure 4:

A. Flow analysis pathway

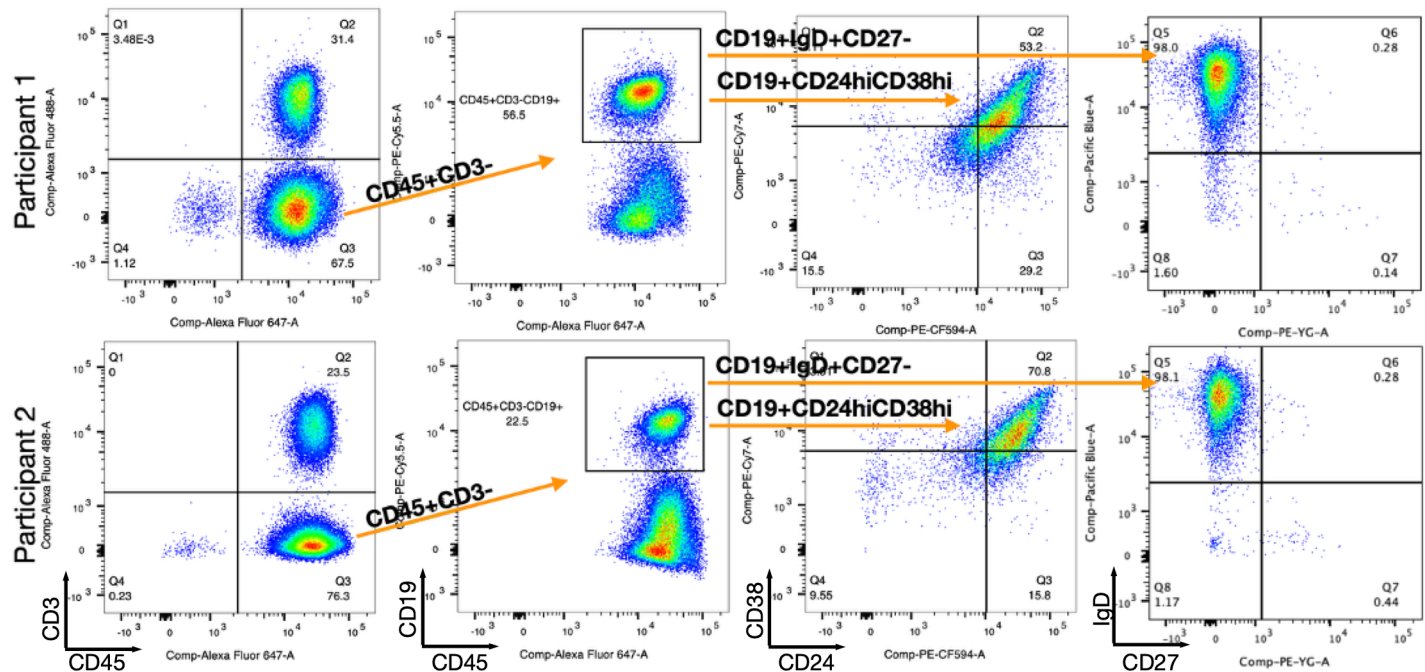

B.

Participant 1

Participant 2

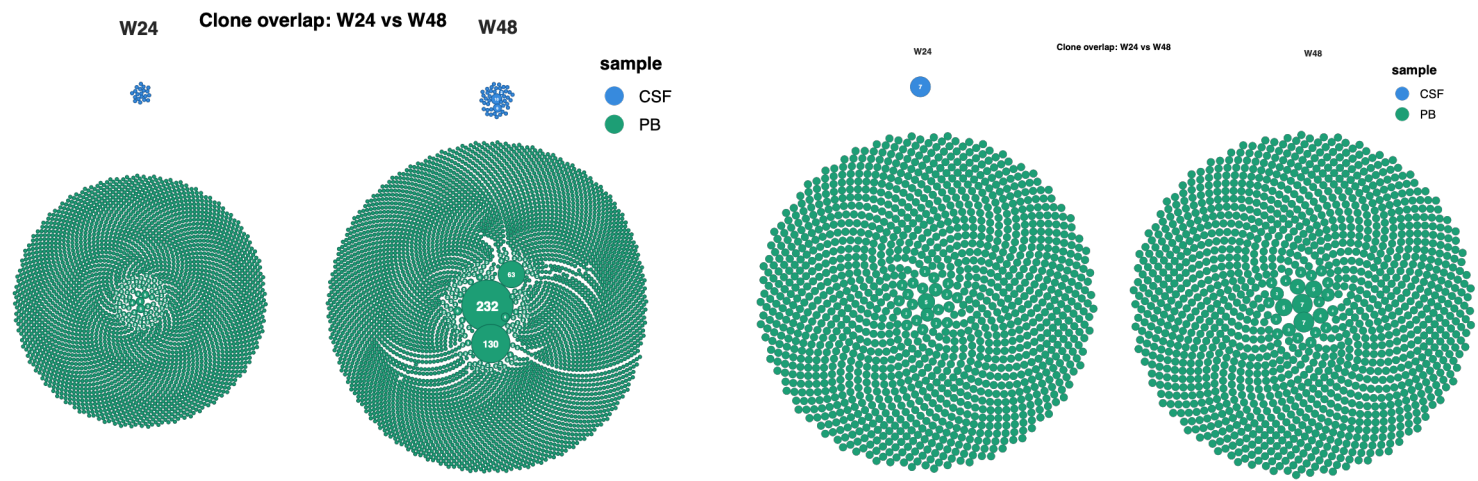

C. CDRH3 length distribution by sample

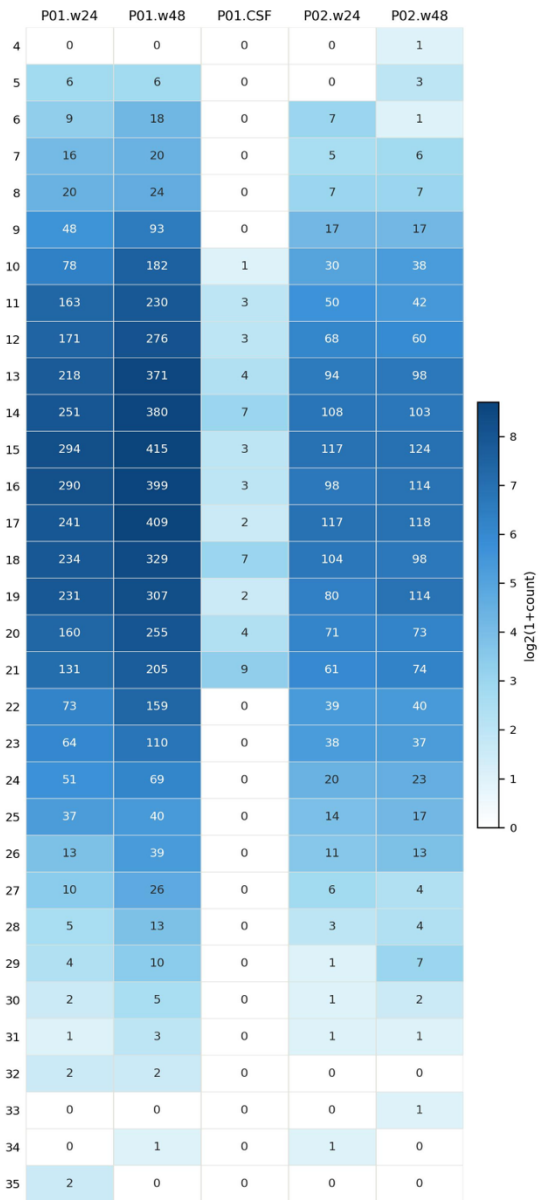

CDRL3 length distribution by sample

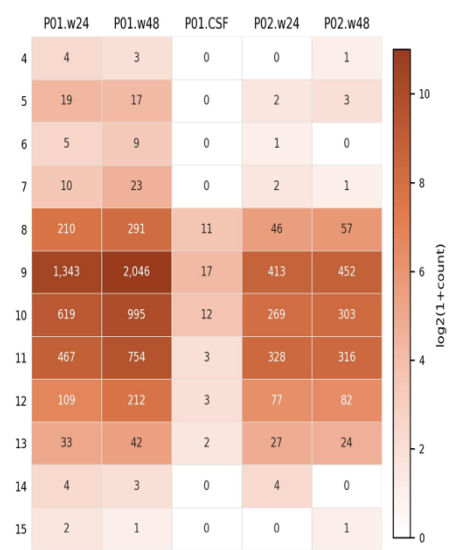

D.

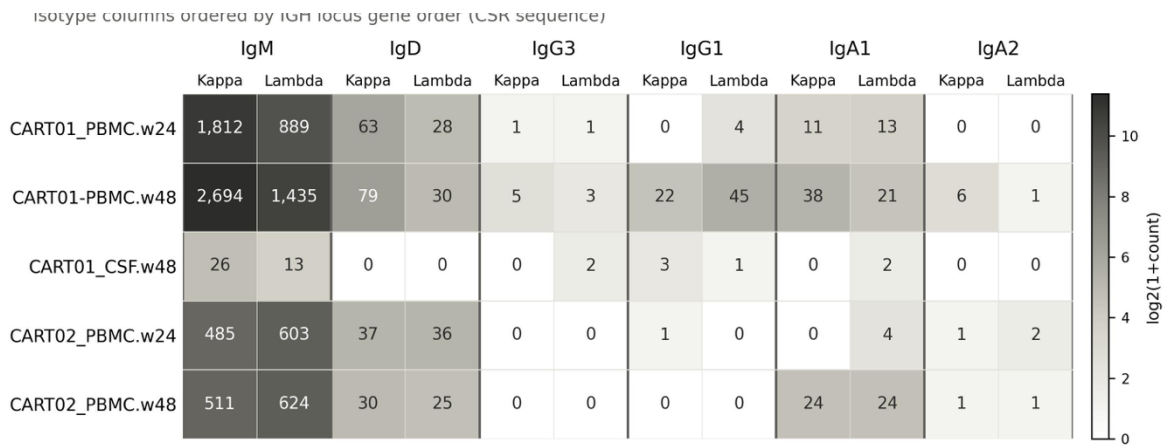

E.

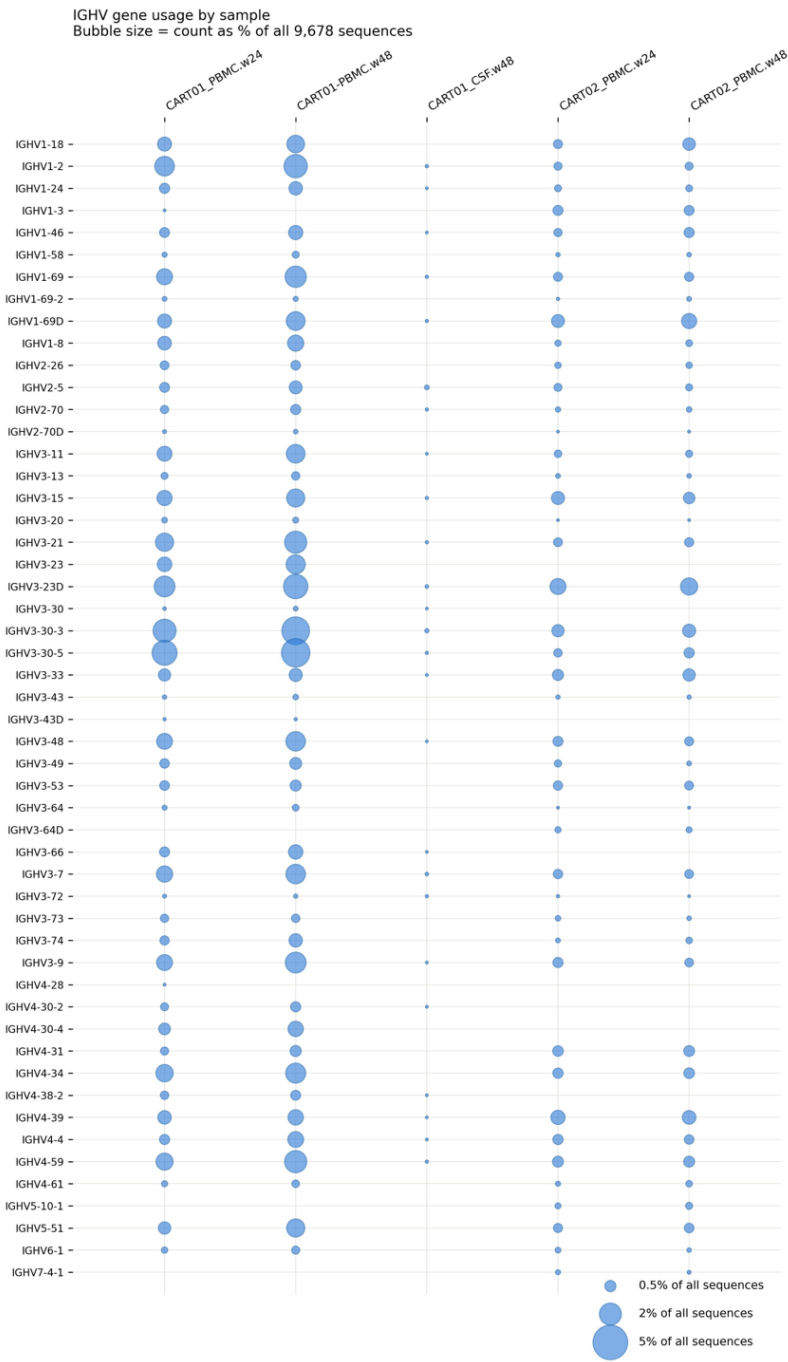

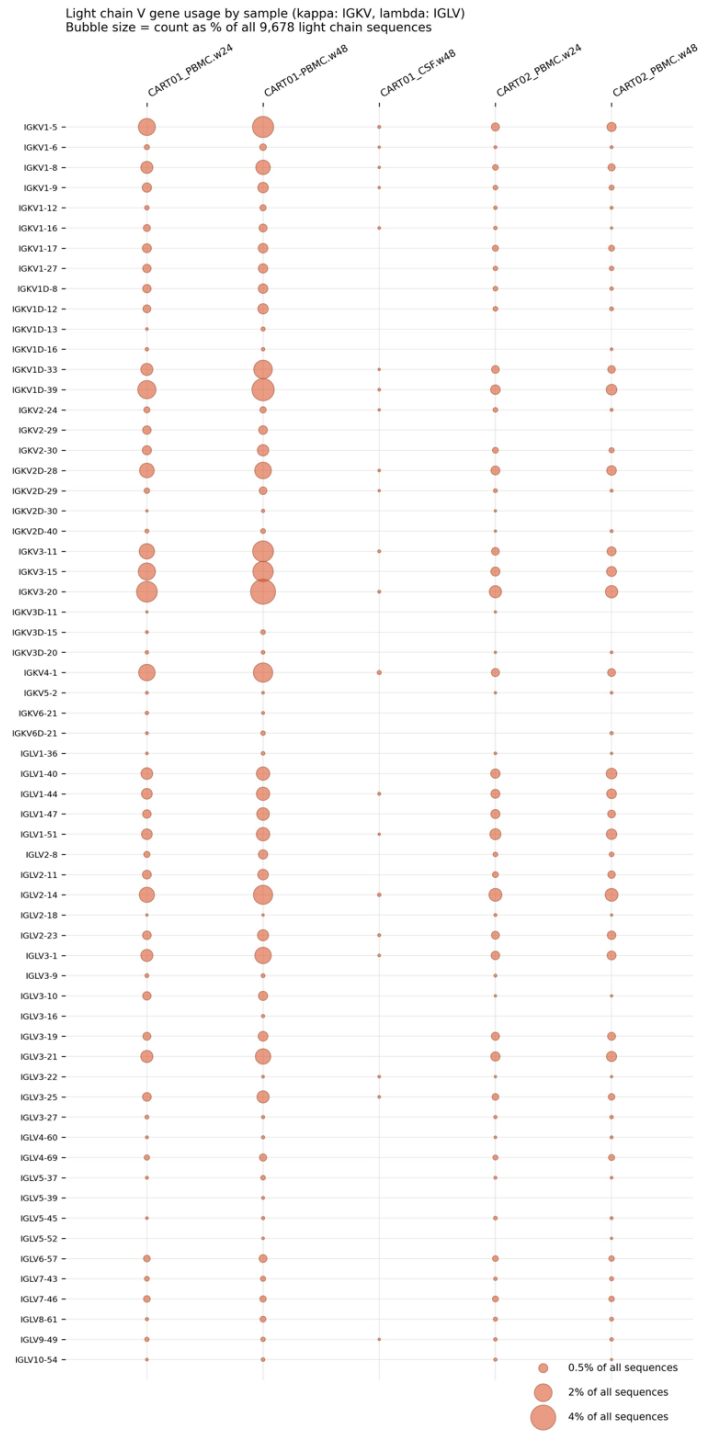

Supplemental Figure 5:

A. B cell activation and homeostasis

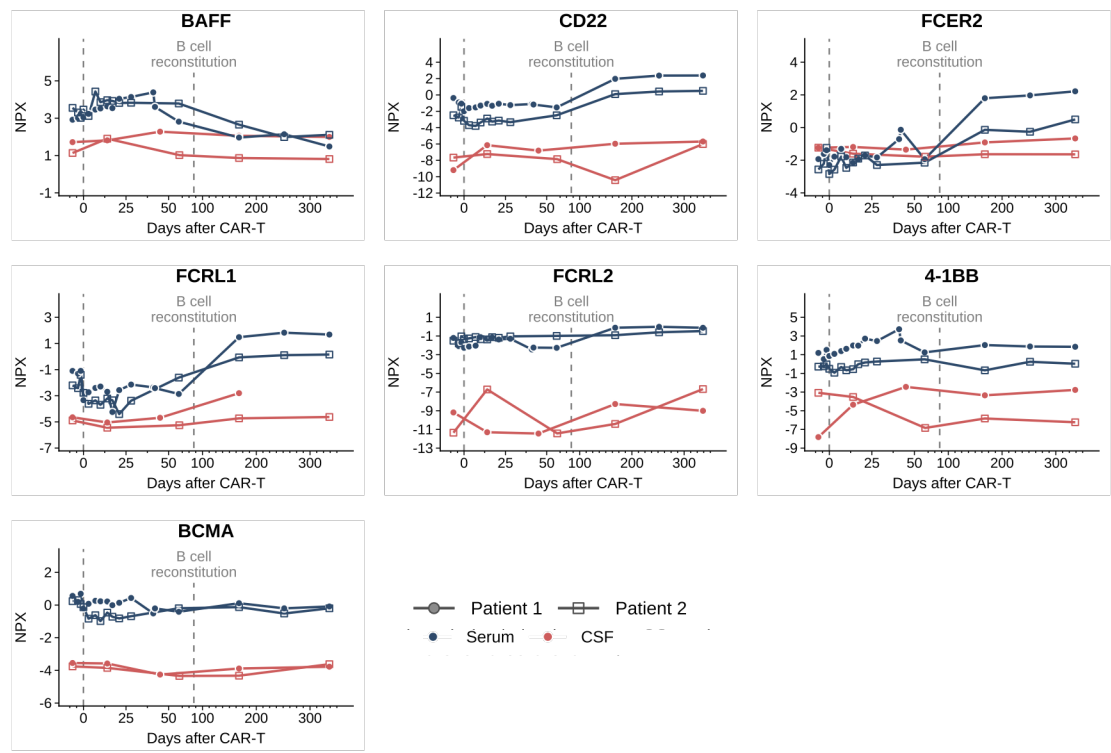

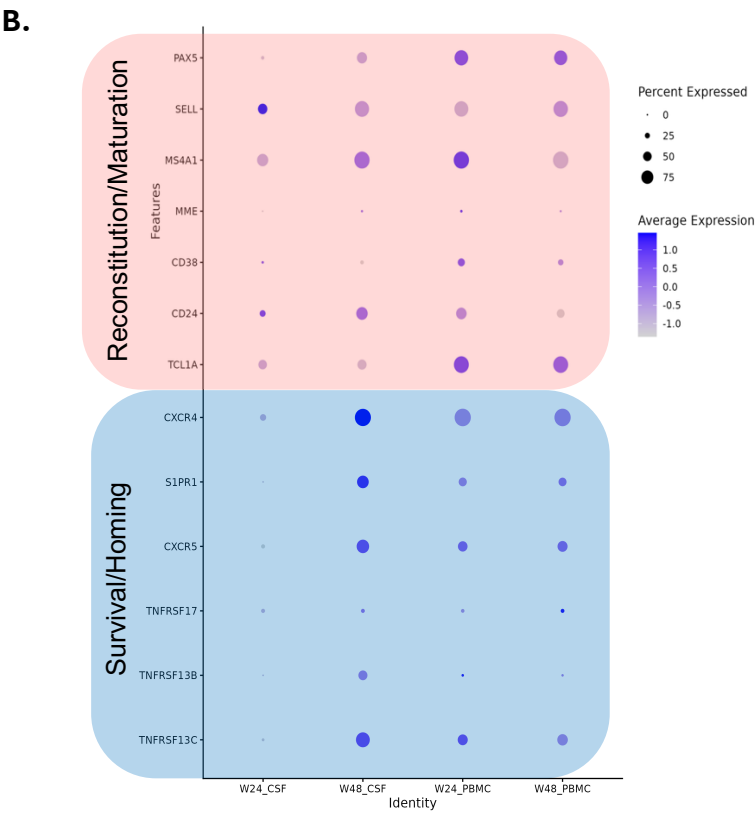

Supplemental Figure 6:

A. UMAPs for both participants over time

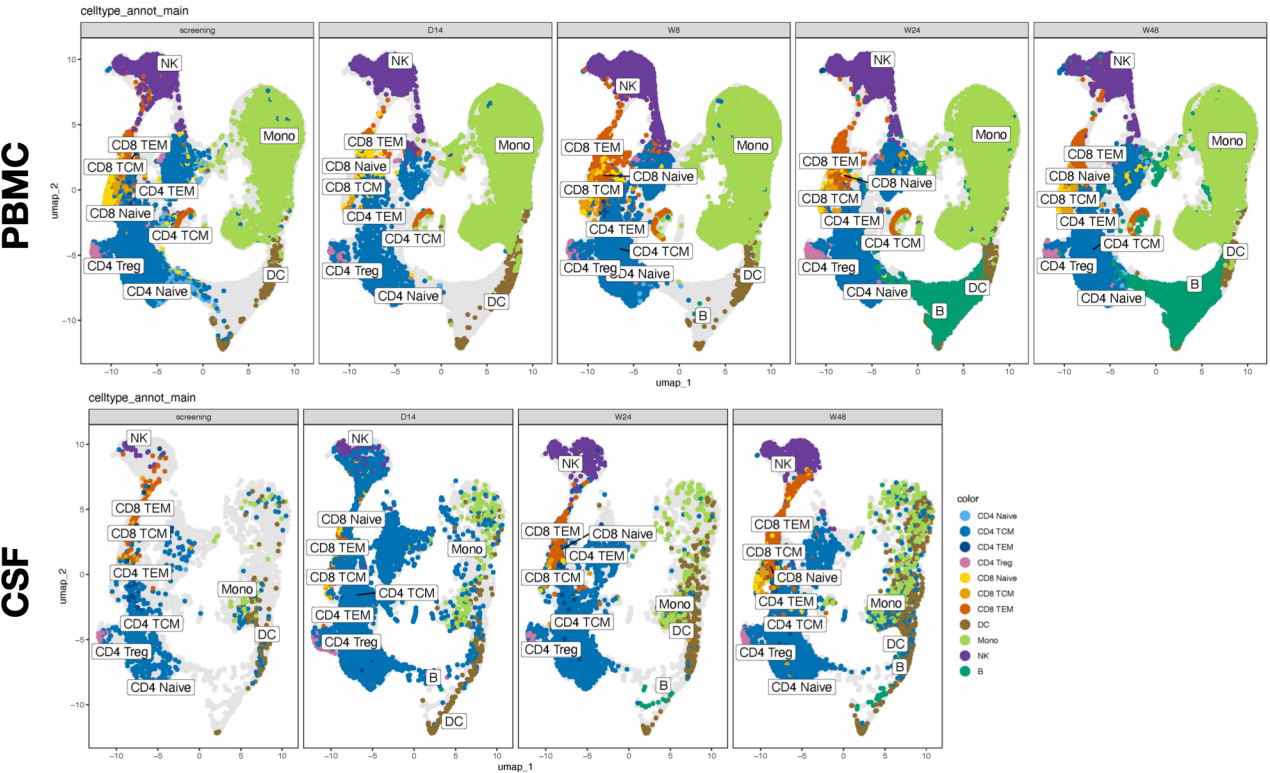

B.

Participant 1

Participant 2

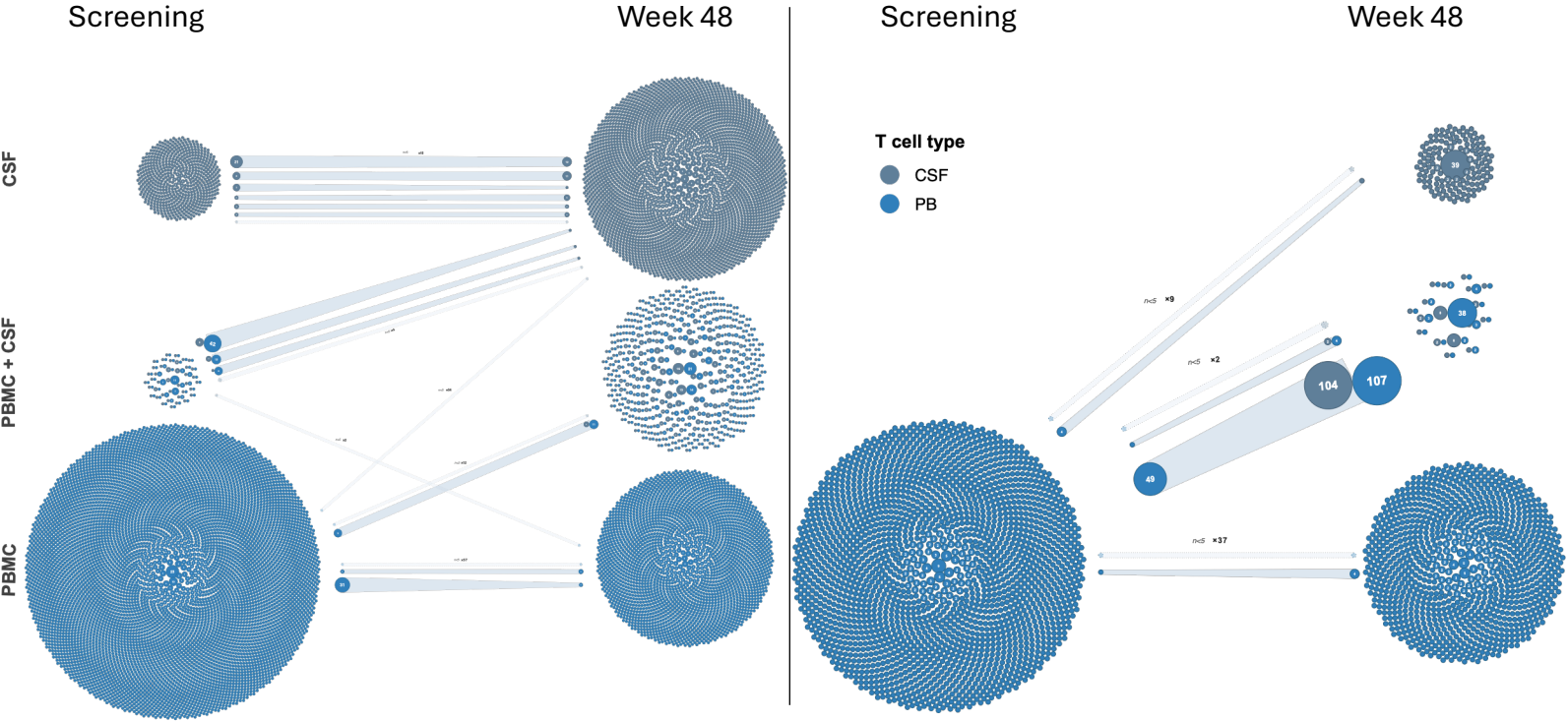

C. Participant 1: all T cells

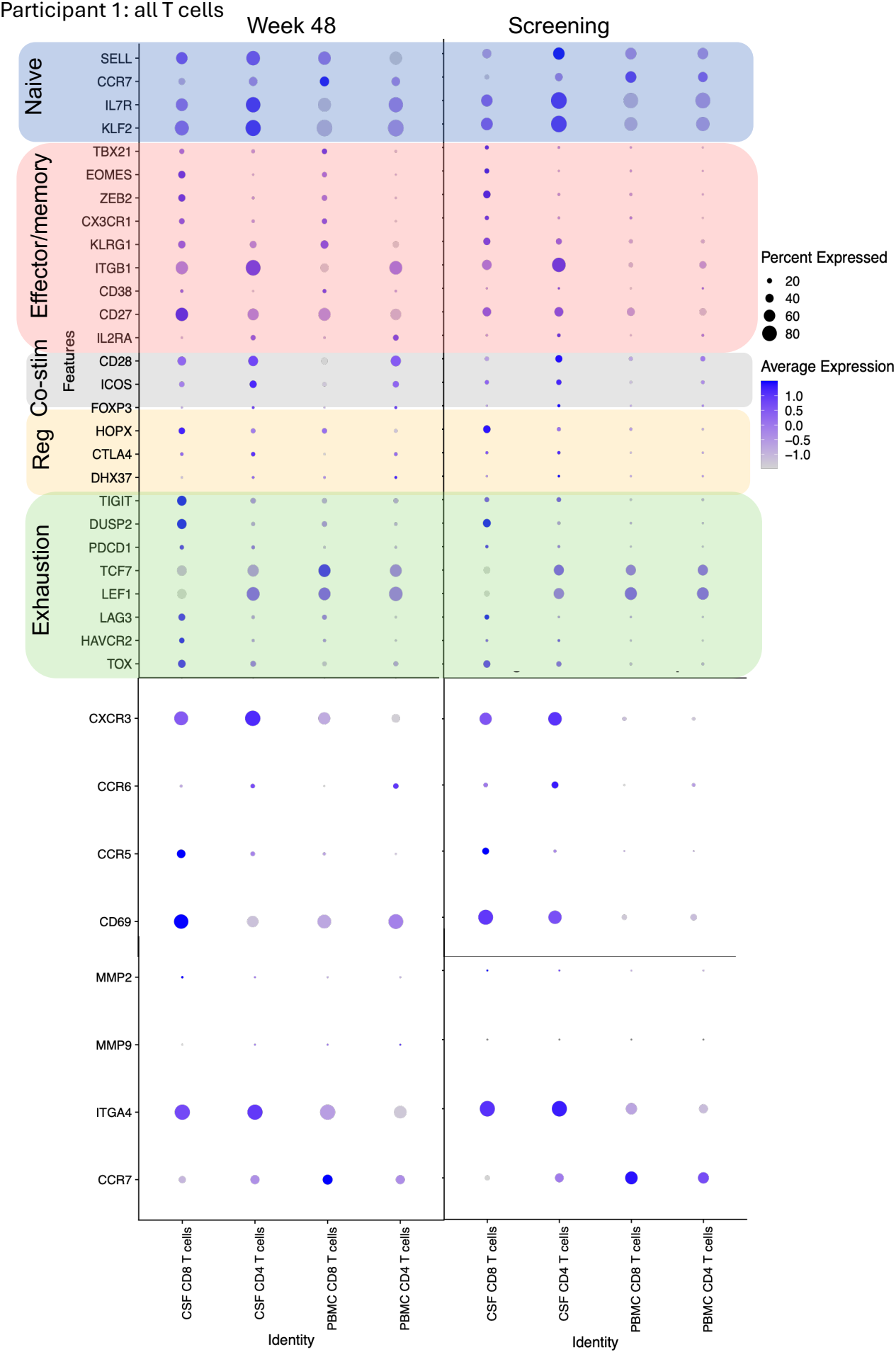

D. Participant 2 (no screening CSF): all T cells

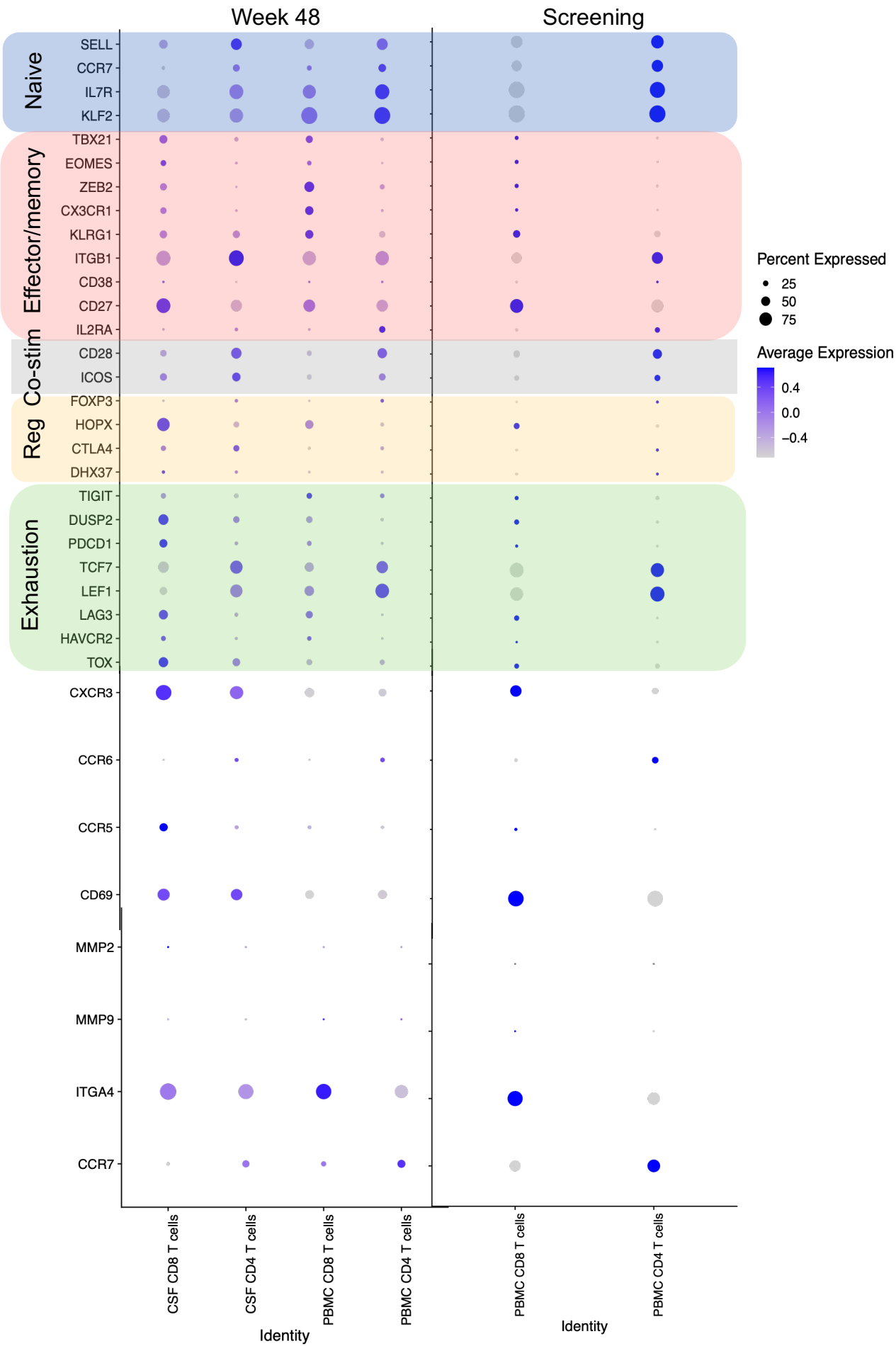

E. T Follicular Helper Cell Genes, all T cells

Participant 1:

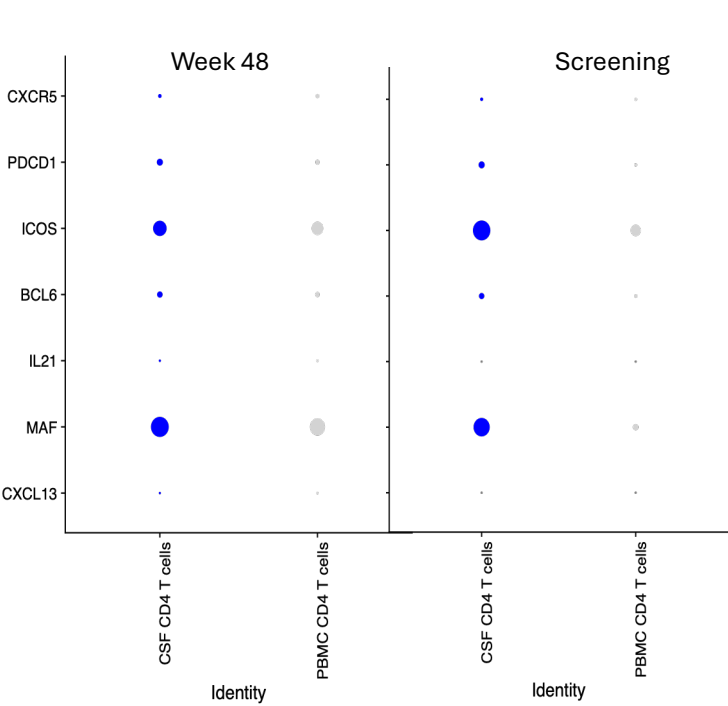

Participant 2:

F. Analysis of viral T Cell Receptors (TCRs) over time.

—

## 1.

### Microglia and myeloid activation

### Astrocyte and neuronal injury

### Other markers

● Serum ● CSF ● Patient 1 □ Patient 2

Sample —●— Serum —●— CSF Patient —●— Patient 1 —□— Patient 2

### Supplemental Figure 7:

#### Supplemental Table 1: T cell response of Participant 1's Adverse Event:

On Day 32, the participant experienced minor facial swelling thought to be attributed to seasonal allergies for which she took cetirizine and Benadryl over-the-counter medications. On Day 34, she developed a sinus headache that resolved with Excedrin. On Day 35, the facial swelling persisted, and the participant noted a new pruritic rash on her neck and chest. On Day 38 a new grade 3 transaminitis was noted and the absolute lymphocyte count (ALC) increased from $0.26$ to $1.09 \times 10^9/L$ as compared to the preceding value on Day 32. On Day 41 the rash progressed and the participant was admitted on Day 42. An abdominal ultrasound revealed mild splenomegaly. Given the rise in ALC we wanted to ensure a thorough investigation was done to understand the underlying biology of what occurred during this time. Based on clinical lymphocyte subsets the ALC was T cells driven with a rise in CD8 cells from $50$ to $309 \times 10^6/L$ and a rise in CD4 cell increase from $41$ to $142 \times 10^6/L$ with no significant change in CD19 cells ($<20 \times 10^6/L$) and a slight decline in NK cells ($120$ to $79 \times 10^6/L$) within 14 days. In looking at the clinical T cell breakdowns, the CD3+ cells were noted to be CD45RO+ suggesting an activated memory phenotype. Clinical cytokine panels also showed elevation in IL-1 beta, IL-2, and soluble IL-2 receptor suggestive of a T cell expansion. Surprisingly IL-10 was also elevated. IL-6 was normal despite a slight elevation during CAR-T expansion. Proteomic analysis also supported a T cell response with upregulation of IL-15, IL-2, IL-7, and TNF alpha (Supplemental Figure 6I). In addition, we observed a marked elevation of serum IL-5, a key eosinophil-recruiting cytokine implicated in drug reaction with eosinophilia and systemic symptoms (DRESS) syndrome [79], alongside evidence of myeloid activation and tissue injury reflected by increased CHIT1, CHI3L1, CSF1R, and TREM2 levels.

There was a notable rise in CD8 cells in both the peripheral blood, rising from 20% to 28%, and in CSF from 20% to 50% in the non-CAR-T positive T cells (Supplemental Figure 2B). On single cell analysis there were 6 CAR-T positive cells found (Supplemental Figure 2B), all of which were CD4 and 5 of 6 were TCM cells. Based on TCR repertoire there was no clear clonal expansion, but a polyclonal non-CAR-T response that was not present at D14 (Supplemental Figure 2C). Gene expression revealed these T cells were mainly CD4 TCM and CD8 TEM consistent with the clinical T cell subtypes. There was further decline in T regulatory cells (Supplemental Figure 2B). Overall, this supports an endogenous proliferative non-CAR-T cell response.
